# USFDA Oncology Drug Approvals from 2014- 2025: Comparing non-immunotherapy and immunotherapy approvals, their endpoints, and their efficacy

**DOI:** 10.64898/2026.10.21.26366185

**Authors:** Neil Bapodra, Katherine G. Morse, Susan E. Bates, Tito Fojo

**Affiliations:** Neil’s current address/institution [MD Anderson Center]; Department of Biochemistry & Molecular Biophysics, Columbia University Irving Medical Center; Department of Medicine, Division of Hematology Oncology, Columbia University; James J. Peters VAMC

**Keywords:** FDA, EMA, Immunotherapy approvals, Non-immunotherapy approvals, Surrogate endpoints, Progression-free survival, Overall survival, Crossover, Censoring

## Abstract

In the United States the Food and Drug Administration (FDA), and in Europe, the European Medicines Agency (EMA) are responsible for regulatory approvals of anti-cancer therapeutics. We examined regulatory approvals by both agencies in the period between 2014 and 2025 and compared immunotherapy and non-immunotherapy approvals for the treatment of cancer. We asked whether the approvals were similar in terms of trial size, endpoints, and development time. We present the data based on the FDA approvals given that the US agency has approved some therapies either rejected by or yet to be approved by the EMA. Beginning with the approval of pembrolizumab for melanoma in 2014 through early 2025 we identified 143 immunotherapy and 211 non-immunotherapy approvals. In the first few years the time to approvals of immunotherapies was markedly less but over time the duration has approached that of non-immunotherapies – with durations of 2079 and 2357 days in 2024/2025 due largely to a median trial size of 461 and 698 participants. Surrogate endpoints have overwhelmed overall survival as the primary endpoint for approval, and we find that with non-immunotherapies only a median of 60% of the benefit predicted from progression/disease/recurrence/event-free survivals translates to overall survival, significantly less than the 99% of the surrogate benefit translated to overall survival for immunotherapies. While the FDA and EMA approval portfolios are very similar, the EMA is on the whole more skeptical/conservative in its approvals. The last decade has seen substantial advances, but much remains to be accomplished against a formidable foe. The size and duration of trials need to be addressed and meaningfully reduced and the validity of surrogates defended.

## INTRODUCTION

The era of immunotherapy was launched with objective cancer regressions observed following the administration of ipilimumab, an antibody against cytotoxic T lymphocyte-associated antigen 4 (CTLA-4) at the time known as MDX-010, in patients with metastatic melanoma^1^. More than a decade later, on March 25, 2011, ipilimumab (Yervoy®) was approved by the United States Food and Drug Administration (FDA) to treat unresectable/metastatic late-stage melanoma. Three years later, September 4, 2014, pembrolizumab received FDA approval, also for melanoma. We report on FDA approvals for the ensuing decade from that date onward, comparing the 143 immunotherapy approvals with the 211 approvals of all other anti-cancer therapies.

Regulatory approval of cancer therapeutics has evolved with a desire to provide patients rapid access to new agents, an acceleration that has been accompanied by substantial debate regarding the strength of the supporting evidence. While traditional FDA approvals required an overall survival advantage, modern oncology approvals are frequently based on relatively small trials, single-arm studies, or surrogate endpoints such as objective response rate (ORR) or progression-free survival (PFS). While advocated to substantially shorten development timelines, surrogate endpoints have often had an imperfect relationship with clinically meaningful outcomes such as overall survival (OS) or quality of life^2–5^. Consequently, many approved therapies enter routine clinical practice despite persistent uncertainty regarding their ultimate clinical benefit. These concerns are particularly relevant for the FDA’s Accelerated Approval pathway, which permits approval based on endpoints deemed “reasonably likely” to predict benefit while requiring subsequent confirmatory trials. However, confirmatory studies have frequently been delayed, have continued to rely on surrogate endpoints, or have failed to demonstrate improvements in OS or quality of life, with many drugs remaining on the market for prolonged periods or converted to regular approval despite limited evidence of definitive clinical benefit.

We describe our analysis of a decade of FDA/EMA regulatory approvals, noting the progress that has occurred but also findings that raise concerns that we feel are addressable and not insurmountable but present challenges^6,7^.

Most of the data is presented as median values and interquartile range [IQR]. We also calculated the median hazard ratio, with IQR, for all trials that reported a hazard ratio (HR) and listed OS or FS as a primary endpoint, respectively.

## METHODS

We conducted a retrospective study of clinical trial endpoints and regulatory timelines for cancer drugs and biologics approved by the FDA and EMA. Data sources included FDA approval letters, Drugs@FDA, ClinicalTrials.gov, EMA assessment reports, and peer-reviewed publications. Approvals were categorized by drug class (immunotherapy vs non-immunotherapy), treatment setting (adjuvant, neoadjuvant, first-line, or later-line), and pathway (regular vs accelerated). For each pivotal trial, we extracted design, cancer type, primary endpoint, sample size, and outcomes. For trials with OS or PFS as the primary endpoint, we calculated absolute differences in median survival between treatment and control arms at approval. We calculated time from first patient enrollment to FDA approval and compared median times for immunotherapy and non-immunotherapy agents. For accelerated approvals, we assessed trial size, duration, and U.S. cancer incidence using SEER or other national databases. We compared FDA and EMA decisions, identifying approvals granted by both agencies or by the FDA alone and examining supporting endpoints and evidence. Finally, we estimated sample sizes required to detect reported hazard ratios with alpha 0.05 and beta 0.20 and compared interim regulatory efficacy data with subsequently published mature outcomes to assess trial efficiency, endpoint robustness, and durability of benefit over time across cancer approvals.

Diseases represented by fewer than three trials were grouped as “Other.” We grouped therapies by treatment class. Immunotherapies were categorized by individual drug, with CAR-T and BiTE therapies grouped as separate classes. Trials evaluating nivolumab plus ipilimumab were counted under both nivolumab and ipilimumab, and trials evaluating durvalumab plus tremelimumab were similarly counted under both agents. Non-immunotherapies were grouped into nine treatment classes. We categorized trials according to whether progression-free survival (PFS), disease-, relapse-event-free survival (DFS/RFS/EFS), overall survival (OS), overall response rate (ORR) DFS/RFS/EFS was listed as a primary endpoint. Trials with more than one applicable primary endpoint were counted in each corresponding endpoint category. We calculated the proportion of trials using each endpoint separately for immunotherapy and non-immunotherapy trials

## RESULTS

We chose September 4, 2014, the approval date for pembrolizumab as our starting point. We assessed the 143 FDA approvals of immunotherapy agents and compared them to the 211 approvals of all other anti-cancer therapeutics, which we refer to as non-immunotherapy agents. Our data harvest continued through the beginning of 2025 before pausing, to allow at least one year to elapse for EMA review and the publication of important data sometimes missing or not yet reached at the time of FDA approvals. Between 2015 and 2025 non-immunotherapies endured median times to approvals after trials were launched of 1352 to 1968 days; varying only slightly from year to year, not surprising for a “class” of drugs with decades of approvals [**Figure 1**]. Immunotherapies on the other hand, were approved at a more rapid pace between 2015 and 2018 with medians of 869 to 1144 days, with durations to approvals eventually increasing to medians of 1338 – 2357 days in the 2019 to 2025 period, often exceeding a time to approval of five years (1825 days) after the launch of a trial. For immunotherapies, the much shorter times to approvals between 2015 and 2018 were often achieved with smaller trials, often a single arm, with a larger percentage of accelerated approvals, and efficacy in the majority assessed wholly or in part by overall response rates (ORRs) and duration of response (DOR). Over time as the most sensitive tumor types were identified and approvals attained, initial enthusiasm over immunotherapy waned and the time for approvals lengthened. Over the decade the difference in time to approval between immunotherapy and non-immunotherapy agents narrowed. Trial size amongst immunotherapies was smaller for BITE’s, Car-T cells and dostarlimab, agents/strategies that often focused on rarer indications but that despite the smaller sizes had comparably long times to approvals [**Figures 2A and 2B**]. A comparison of accelerated and regular approvals with both immunotherapies and non-immunotherapies finds that while accelerated approvals were often granted to smaller trials, their times to approval were similar reflecting the often rarity of diseases in accelerated decisions [**Figure 2C**]. Indeed, across all approvals there was no correlation between the days to approval and size of the trial [**Figure 2D**].

**Figure 1.**
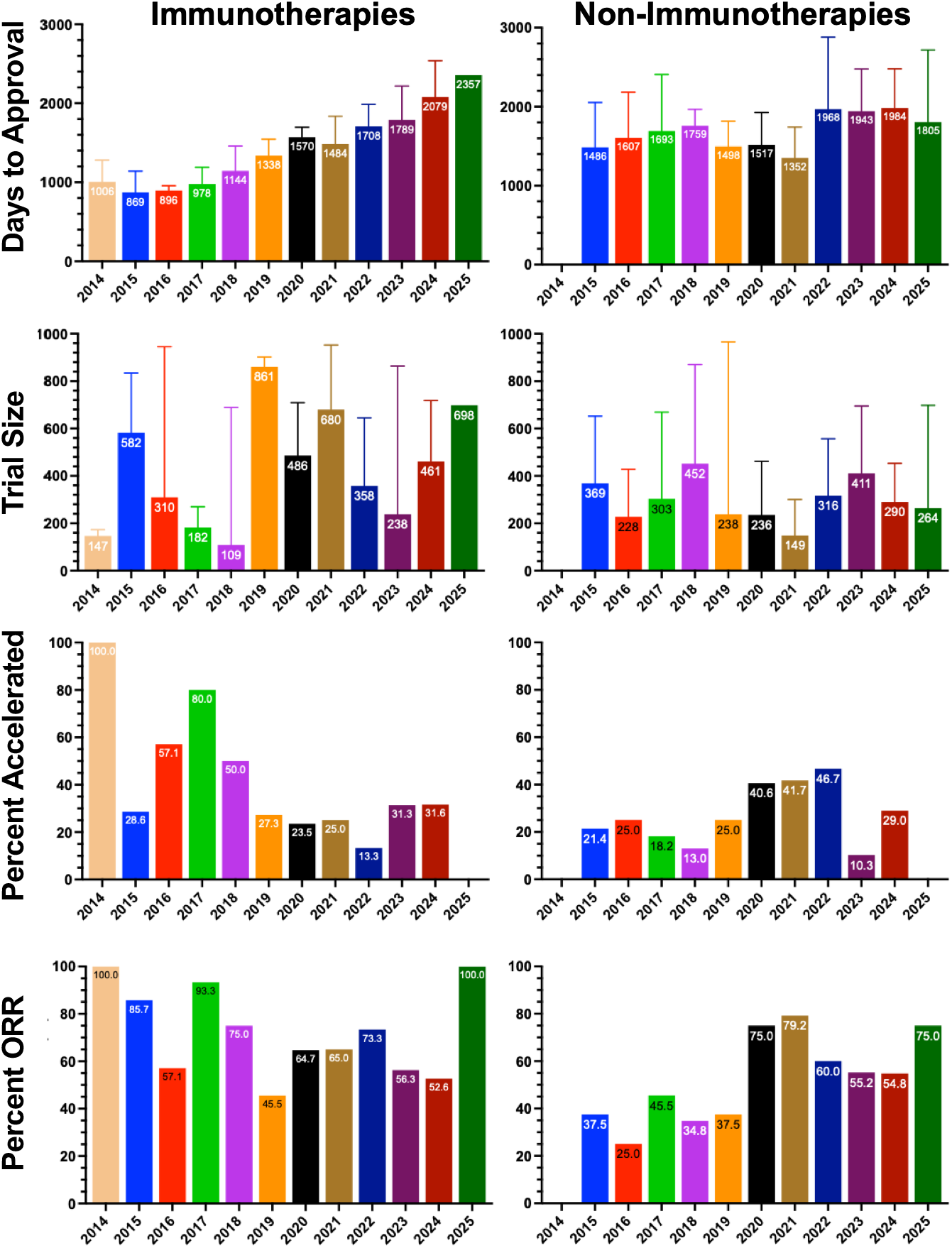
Comparison of immunotherapy and non-immunotherapy approvals from 2014 to 2025. Days to approval, trial size, percent of approvals that were accelerated and percent overall response rate [ORR].

**Figure 2.**
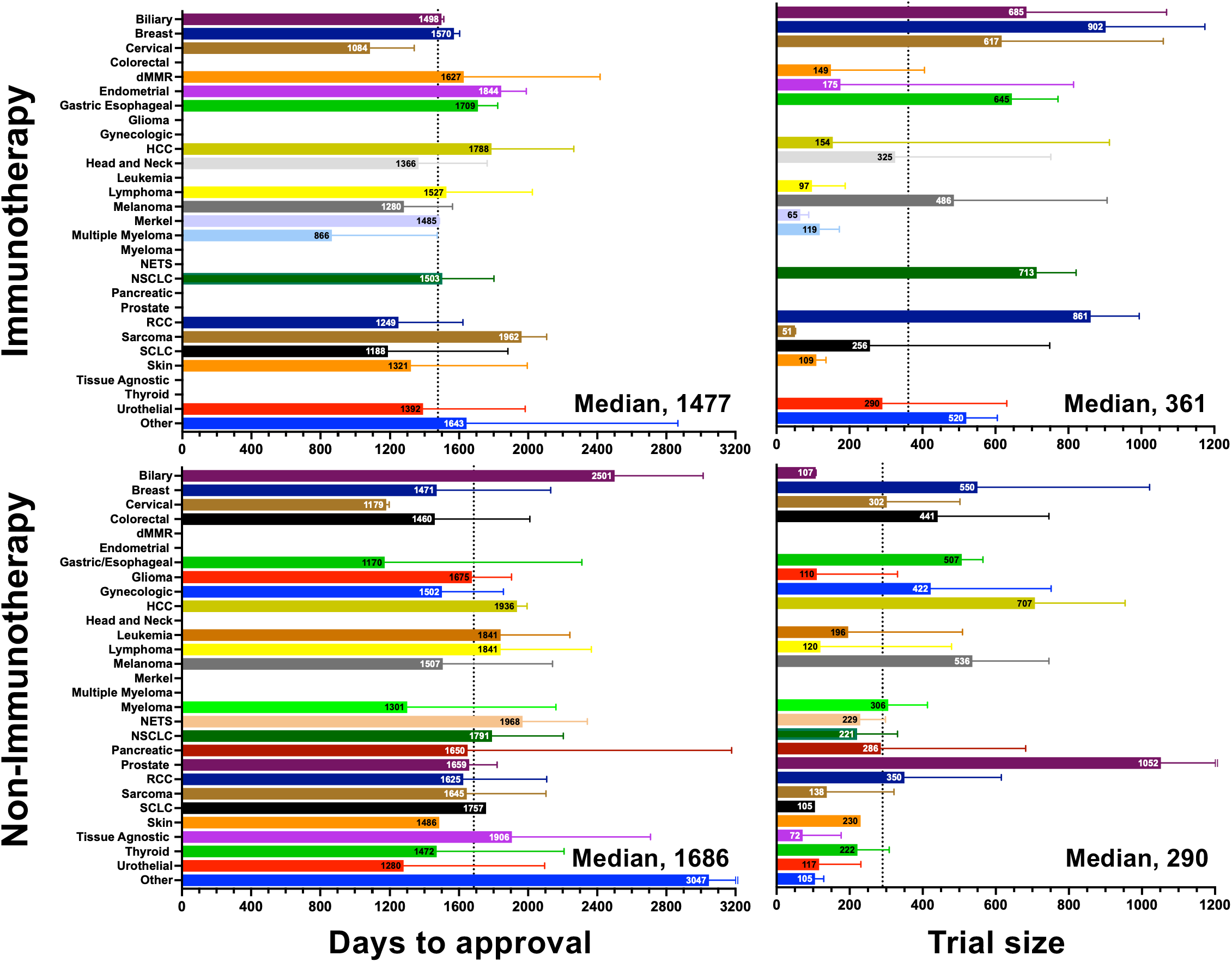
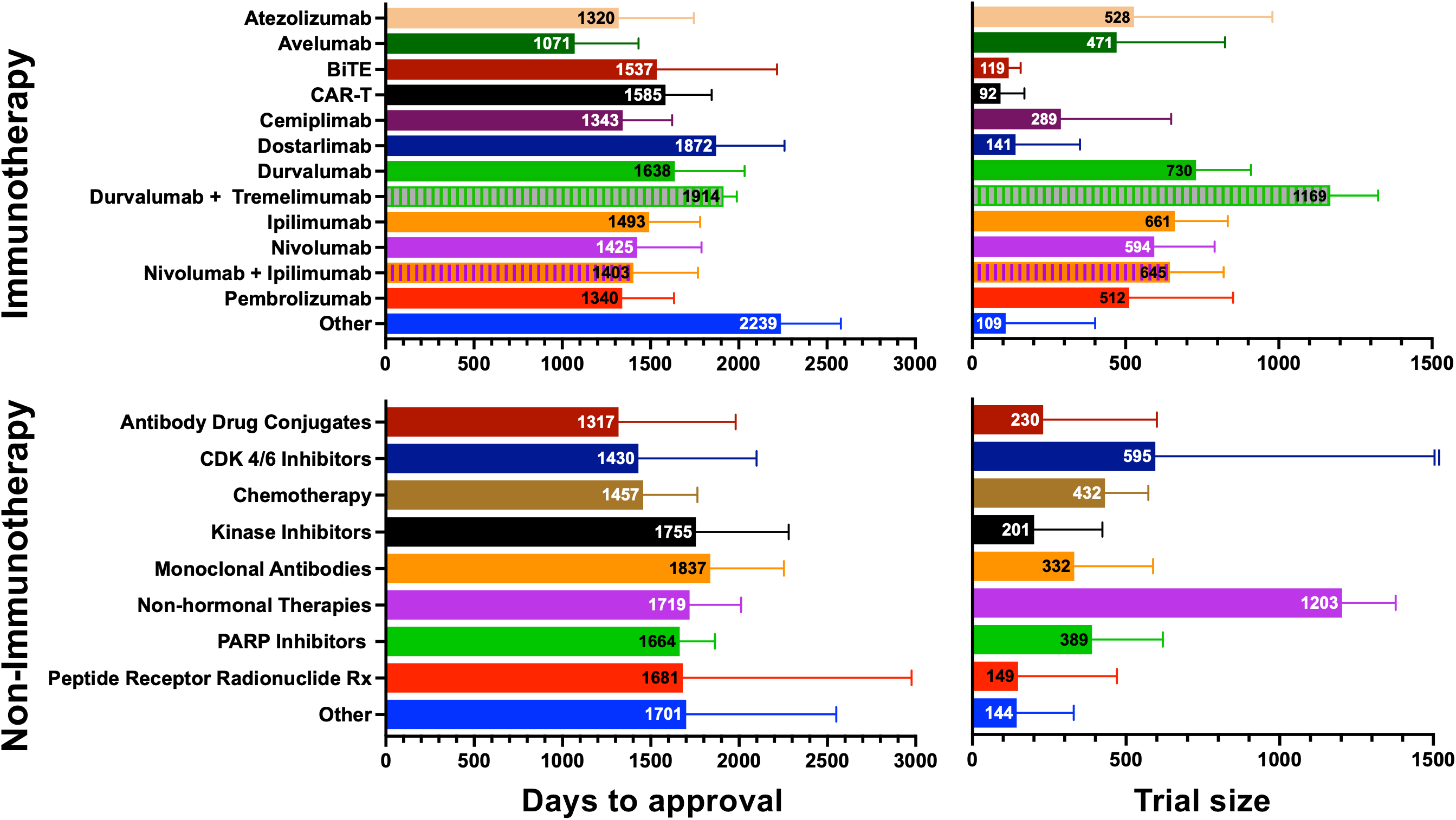
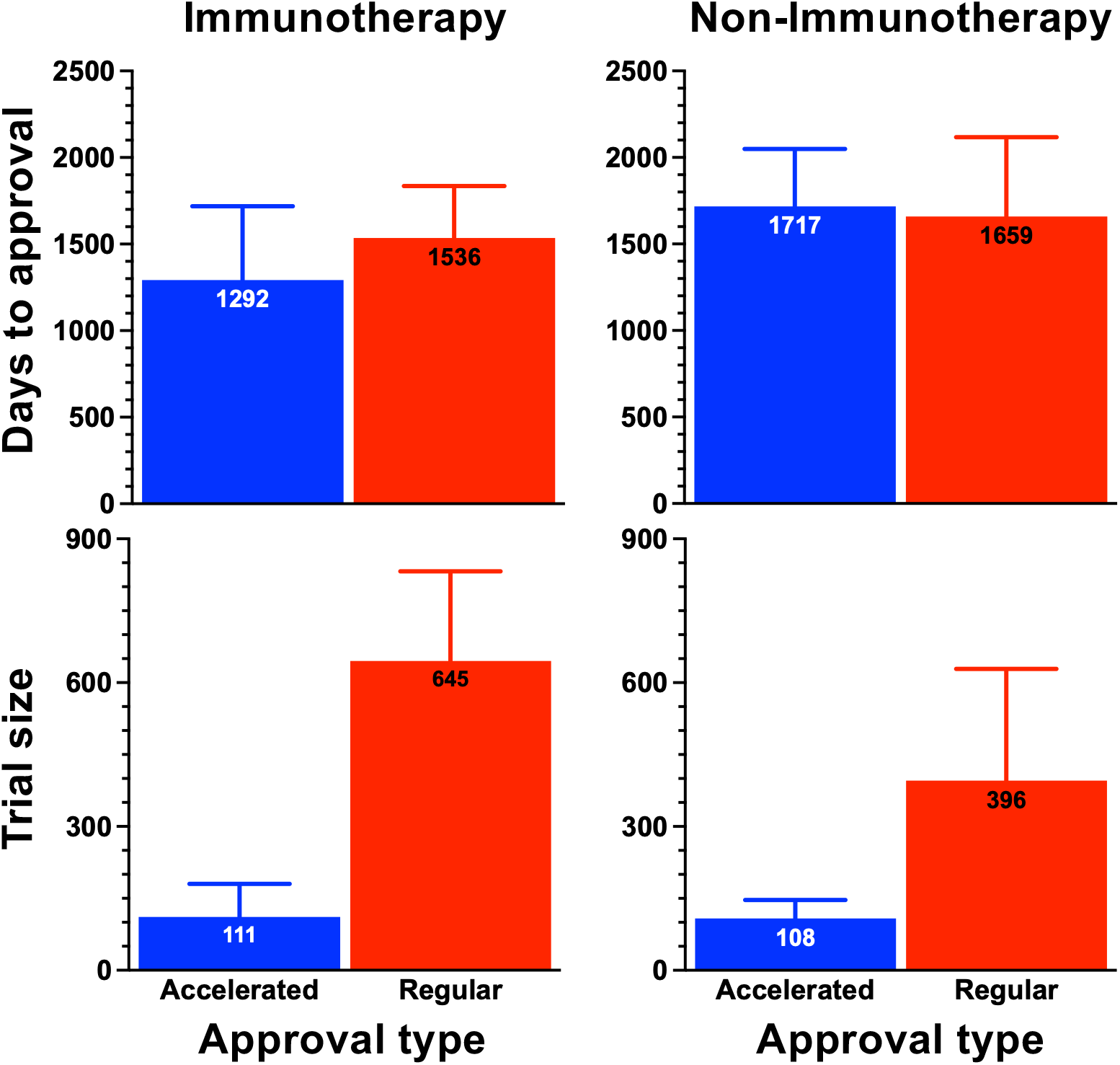
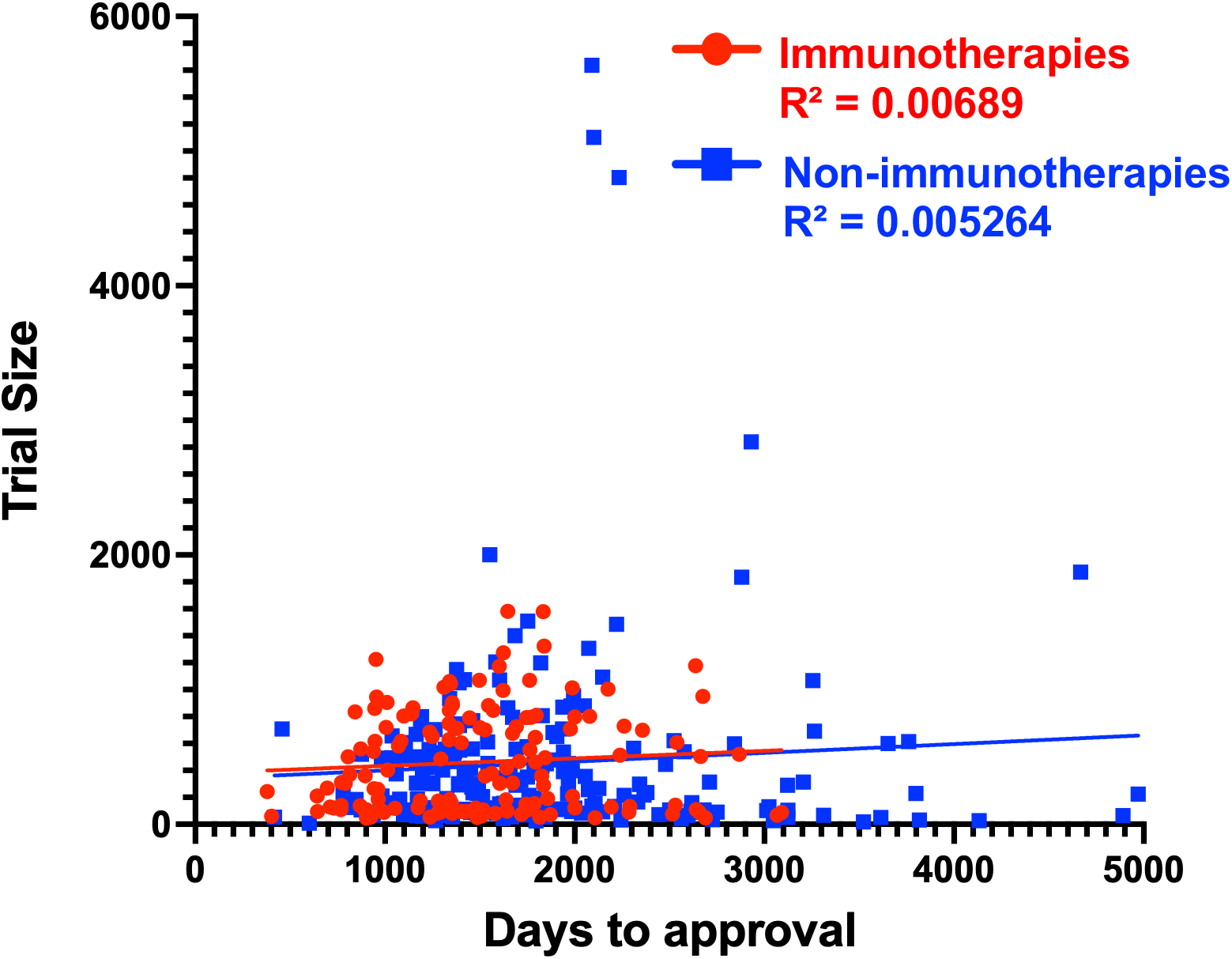
Comparison of immunotherapy and non-immunotherapy approvals from 2014 to 2025. Days to approval and trial size according to: [**A**] the disease investigated; [**B**] the immunotherapy agent or the class of non-immunotherapy drug; [**C**] the approval type (accelerated or regular). [**D**] Days to approval shown to be independent of trial size. In [**A**], diseases represented by fewer than three trials were grouped as “Other.” For immunotherapy, this category included mesothelioma (n=2) and TMB-H tumors (n=1). For non-immunotherapy, it included GIST (n=2), Kaposi sarcoma (n=1), neurofibromatosis (n=2), and neuroblastoma (n=2). Immunotherapies were categorized by individual drug, with CAR-T and BiTE therapies grouped as separate classes. Trials evaluating nivolumab plus ipilimumab were counted under both nivolumab and ipilimumab, and trials evaluating durvalumab plus tremelimumab were similarly counted under both agents. Non-immunotherapies were grouped into nine treatment classes.

Endpoints for assessing efficacy have evolved over time, with “surrogates” replacing overall survival as the approval endpoints, an evolution its proponents have argued would allow trial endpoints to be “read out” quicker. The past decade has seen a marked increase in the use of surrogates with most approval letters citing more than one endpoint as the basis for authorization, often including at least one surrogate. Progression-, disease-, relapse- and event-free survival, (PFS, DFS, RFS, EFS henceforth the “FS endpoints”) together with complete response rate (CR) overall response rate (ORR) and duration of response (DOR), are surrogate endpoints increasingly cited as the evidence for authorization in 74% of approval letters. [**Figure 3A**]. However, surprisingly, the days to regulatory approval for trials where the approval letter referenced a surrogate endpoint as providing support for the authorization was similar to that for immunotherapy trials in which the approval letters cited overall survival as an endpoint with median values of 1498/1573 for immunotherapy and non-immunotherapy trials, respectively, when overall survival as the endpoint and 1477/1699 for immunotherapy and non-immunotherapy trials, respectively with a surrogate as an approval endpoint [**Figure 3B**]. Trial size, on the other hand presented a broader distribution, with trials where ORR and DOR supported the approval letter’s authorization much smaller in size than those with other surrogates providing support, again results impacted by single arm trials in rare disease, taking longer to accrue.

**Figure 3.**
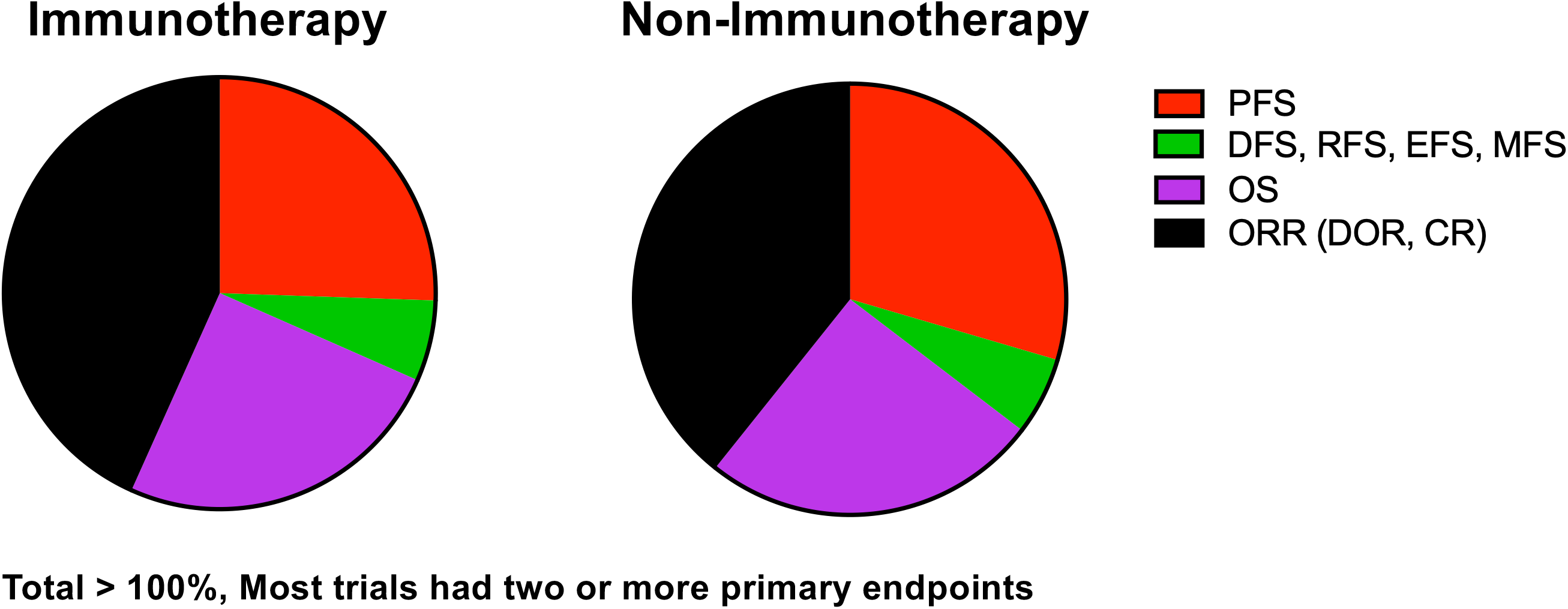
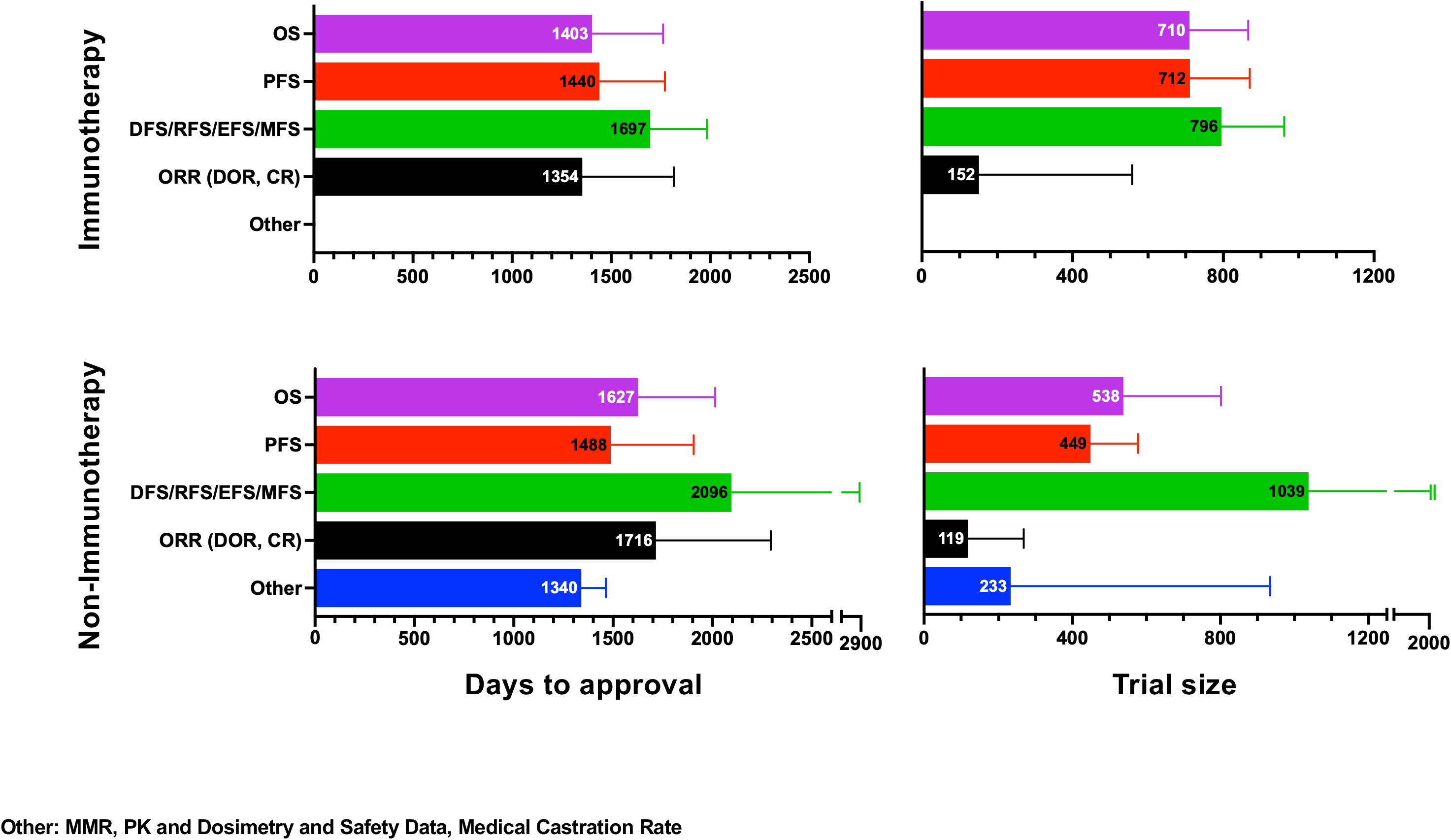
Days to approval and trial size are independent of the approval endpoint. [**A**] Distribution of approval endpoints. Trials with more than one applicable primary endpoint were counted in each corresponding endpoint category. [**B**] Median values for days to approval and trial size for the various approval endpoints.

An important goal of this analysis was to compare the efficacy of approved immunotherapies and non-immunotherapies with a previous analysis published in 2015 that found 2.1- and 2.6-months improvements in PFS and OS, respectively, in non- immunotherapies [Fojo, 2014]. Efficacy can be assessed by examining the gains in median values for the “FS” and OS endpoints and/or by looking to the hazard ratios observed in the trials. We examined the FS and OS gains achieved with non- immunotherapies [**Figures 4A and 4B]** and immunotherapies [**Figures 4C and 4D]** with all summarized in **Figure 5**. Note that the immunotherapy trials often comprised a combination with chemotherapy. Each panel of **Figure 4** is anchored by their left-most panel representing the gains in months achieved in the FS or OS endpoints by the experimental over the control arms of the trial. Especially with immunotherapy, as shown in **Figures 4C and 4D** one can see a trend to higher gains in months with increasing FS and OS in the experimental immunotherapy arm [group of bars second from left], something one might think is not surprising, given there was more “headroom for gains”. Direct comparisons are difficult for a variety of reasons, but qualitative changes can be discerned. With non-immunotherapies [**Figures 4A and 4B]** fewer approvals with FS and OS as their endpoints were first-line trials, but as can be seen in the FS graphs, first-line therapies cluster in the upper part of the graph having achieved the largest gains, an outcome that would be expected. In contrast, the development of immunotherapies [**Figures 4C and 4D**] quickly moved to randomized first line trials and as the graphs show, first-line therapies comprise more than 45% of the immunotherapy approvals based on the FS and OS endpoints. Interestingly the median FS gains with non-immunotherapies at 4.7 months [**Figure 4A**] exceeded the gains of the largely first-line immunotherapy entries at 2.6 months [**Figure 4C**] – an observation at odds with the thought that first-line interventions allow for greater gains. However, direct comparisons are difficult and with the non-immunotherapies these higher values were driven in part by redundant/me-too trials with targeted therapies previously successful in mutation-driven NSCLC, prostate cancer vulnerable to androgen-receptor pathway inhibitors (ARPIs) and studies in breast cancer with CDK4/6 inhibitors and those targeting HER2. Moving further to the right we come to hazard ratios, which were for the most part monotonous at values close to 0.7, increasingly accepted as an approval threshold, although especially in the FS endpoints [**Figures 4A and 4C**] a trend can be seen to lower hazard ratios in the trials at the top, those being ones that achieved the largest gains in median values. Finally, the size of the trials had no correlation with outcomes but was larger in trials with immunotherapies than with non-immunotherapies.

**Figure 4.**
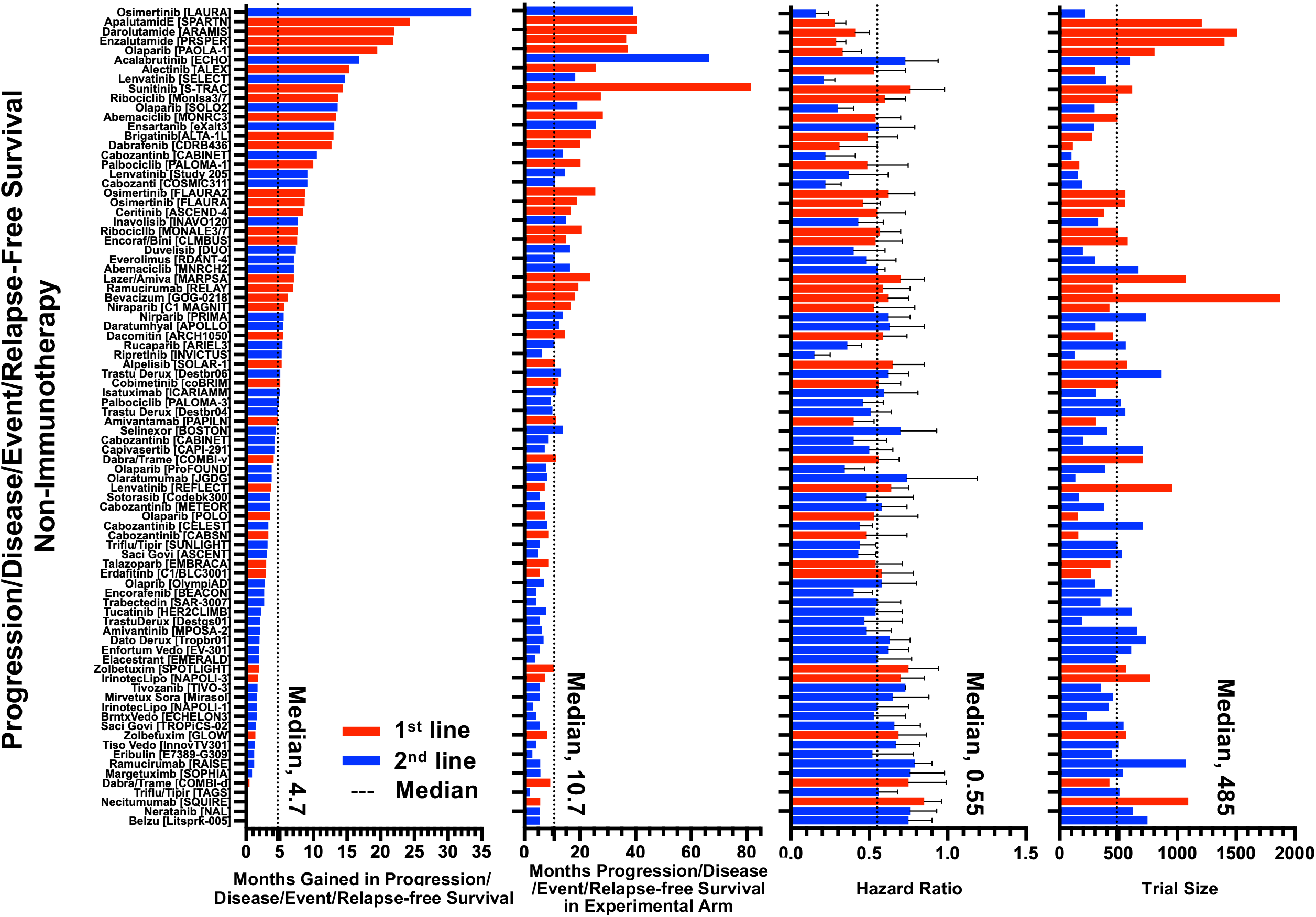
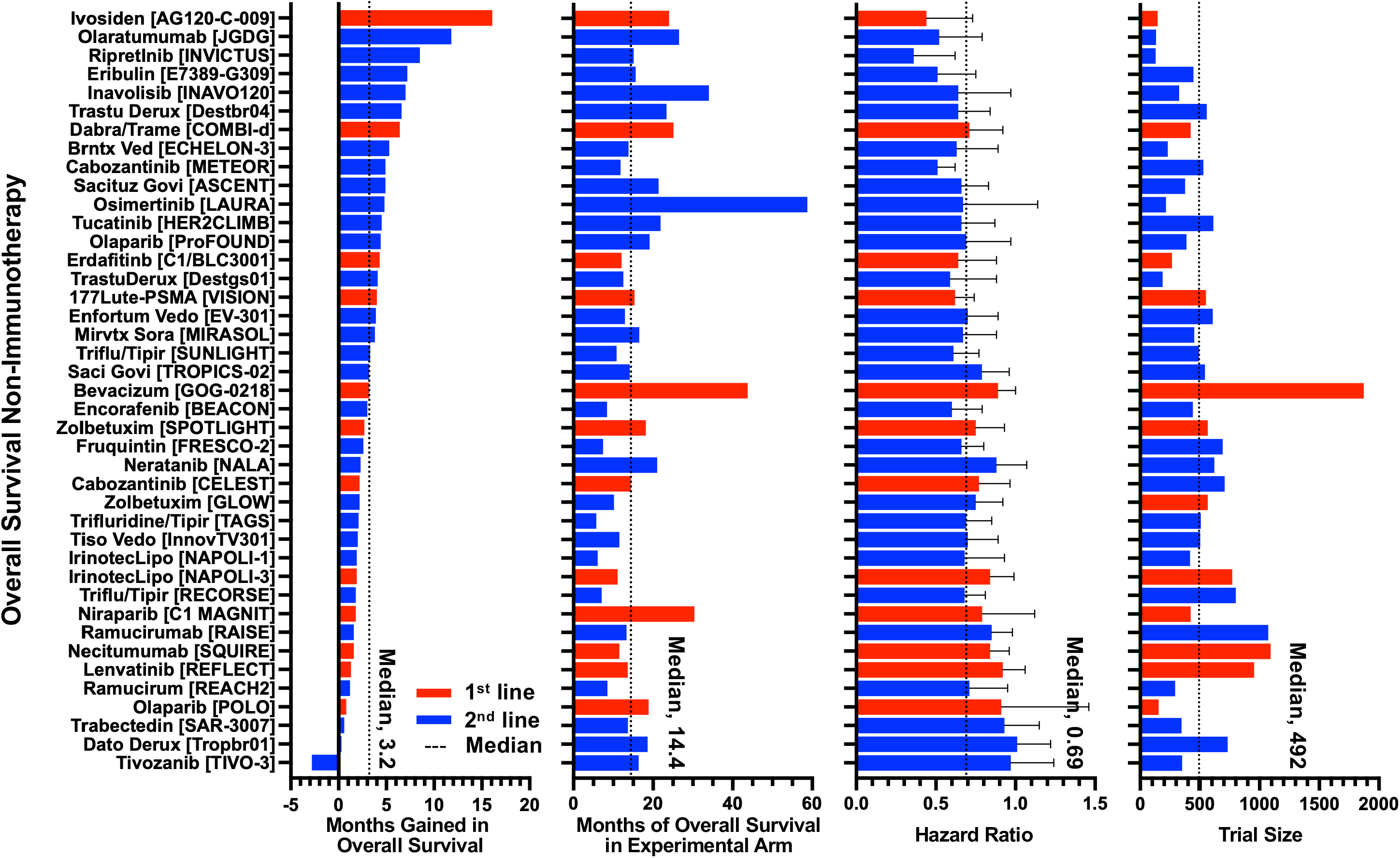
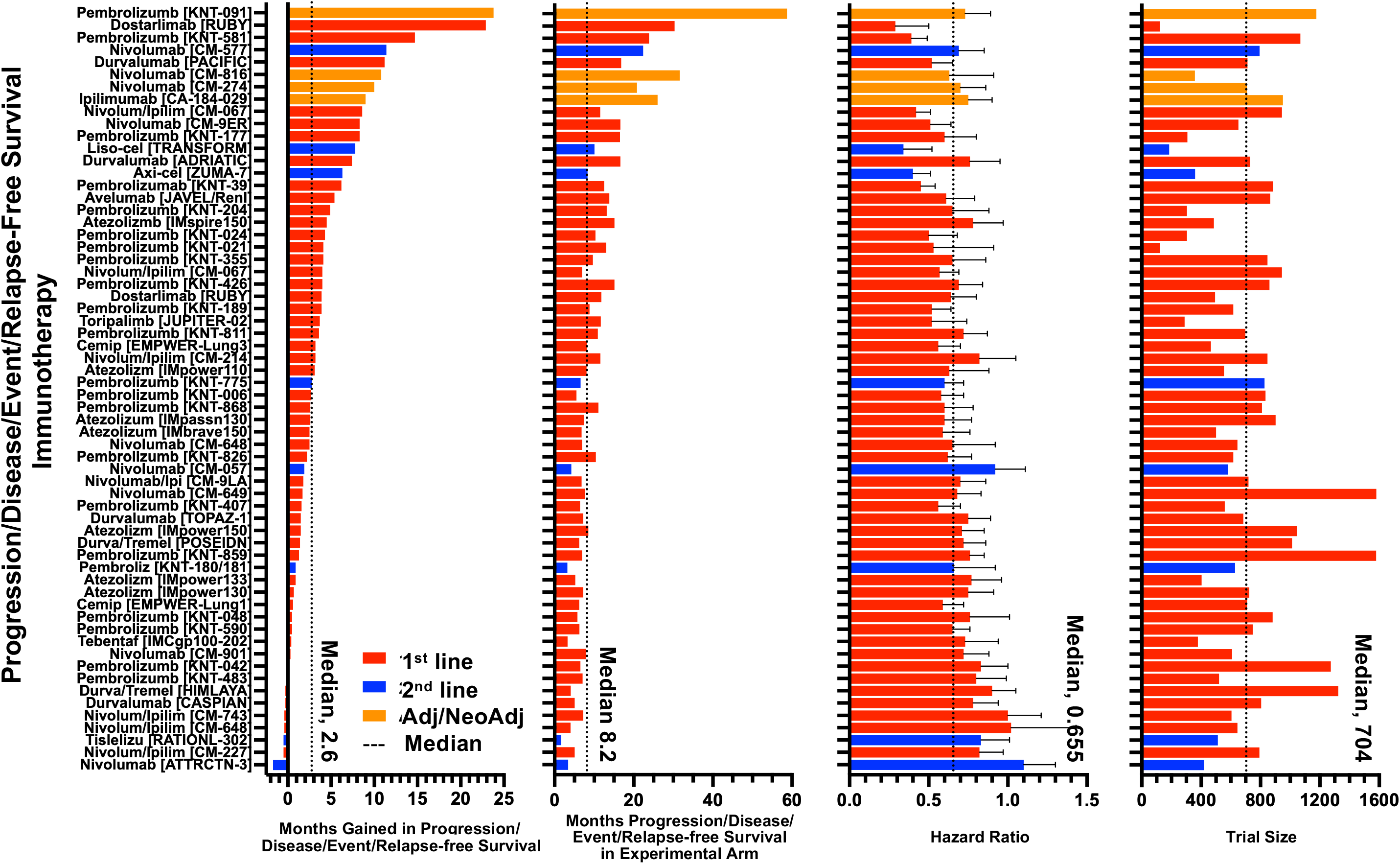
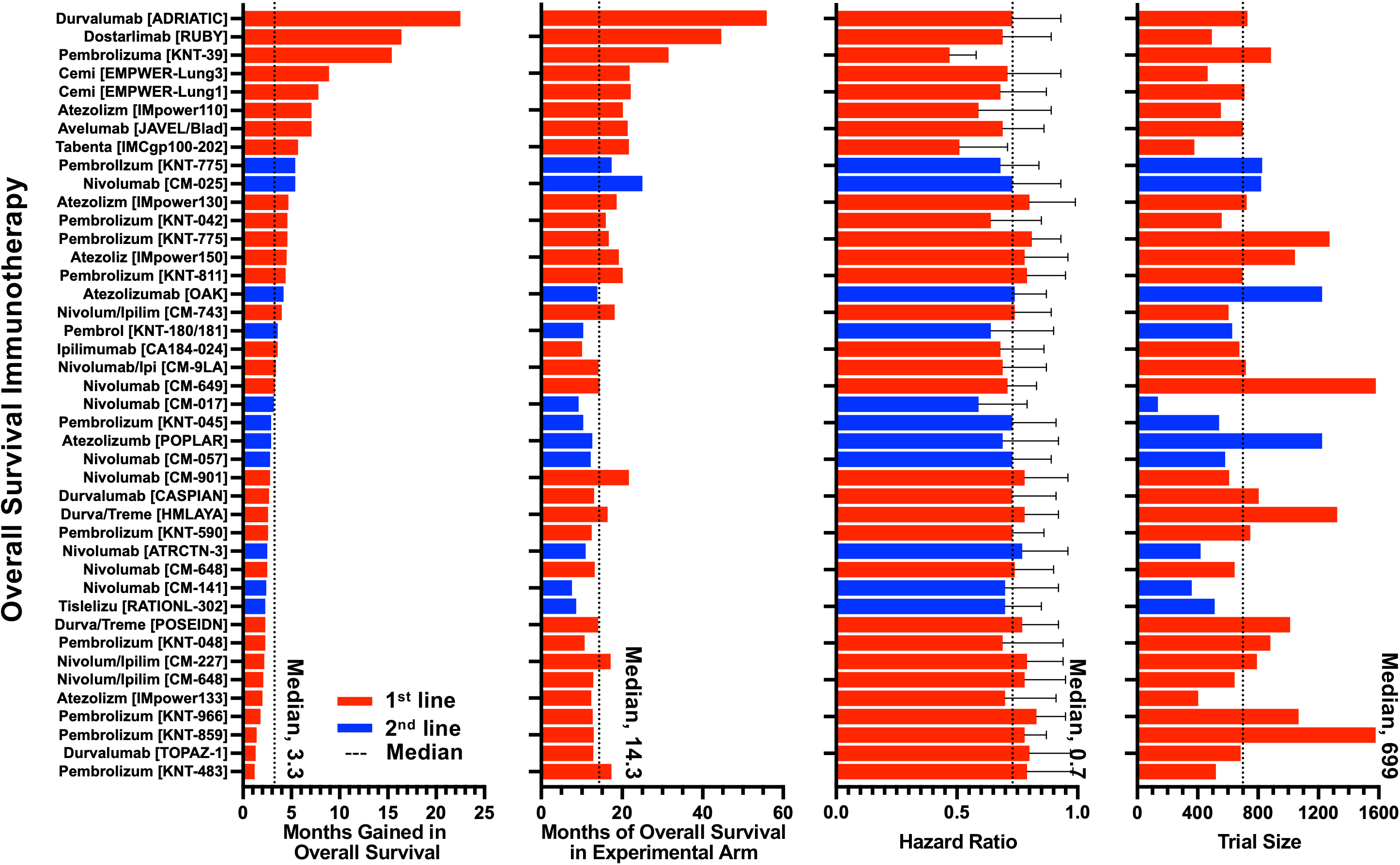
Efficacy assessed by examining the gains in median values for the “FS” (Progression/Disease/Event/Relapse-free survival) and OS endpoints and by looking to the hazard ratios observed in the trials. [**A and B]** FS and OS gains achieved with non-immunotherapies [**C and D]** FS and OS gains achieved with immunotherapies. For each panel from left to right: months gained by the experimental arm in FS or OS [FS/OS Experimental – FS/OS Reference arm]; followed by months of FS or OS in the experimental arm; followed by hazard ratio; and finally, trial size.

**Figure 5.**
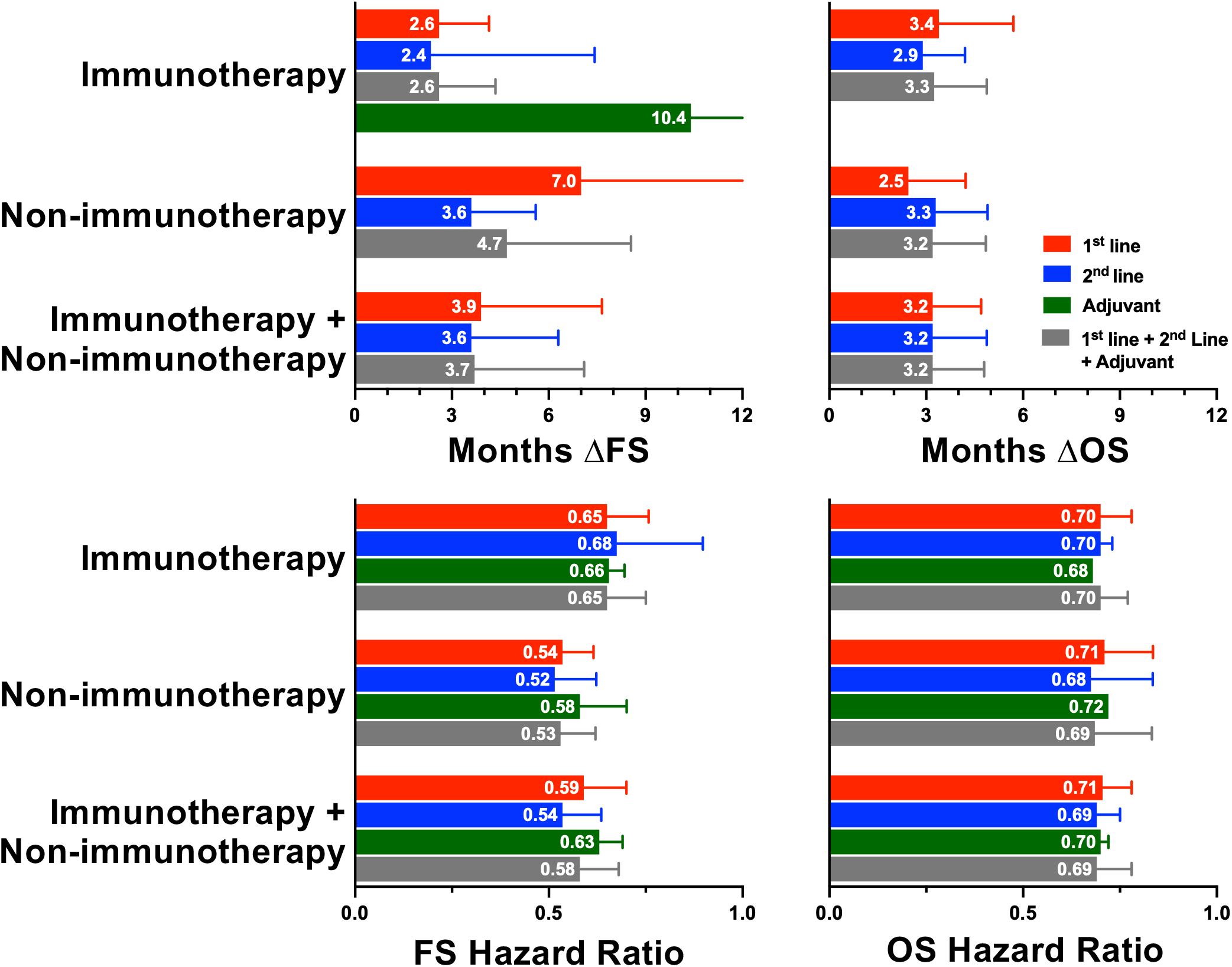
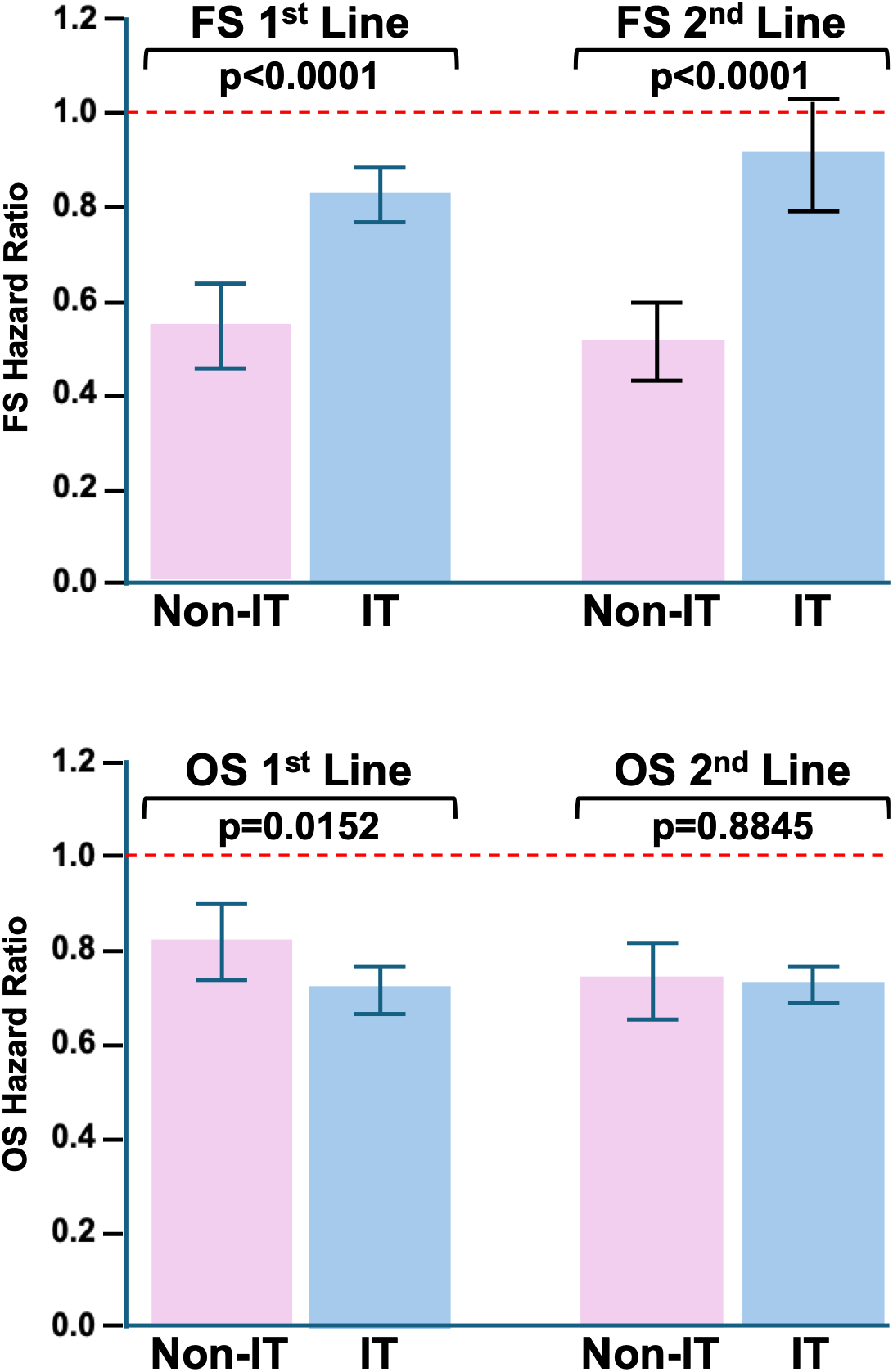
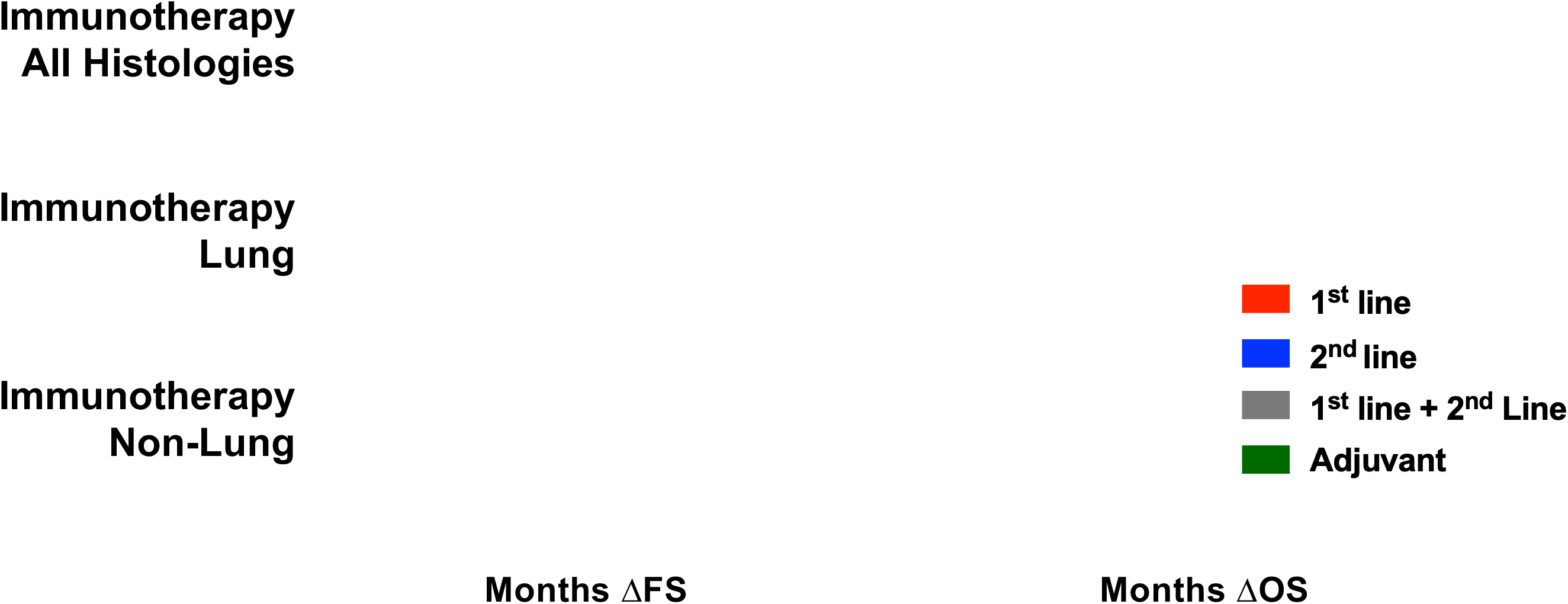
[**A**] Comparison of months gained in FS [Months ΔFS] FS and OS [Months ΔOS] by the experimental arm [FS/OS Experimental – FS/OS Reference arm]; and FS and OS hazard ratios. [**B**] Analysis showing difference between immunotherapies and non-immunotherapies in the FS but not the OS endpoint. [**C**] With differences between the FS and OS endpoints most pronounced in lung cancer. We included only trials that listed OS or FS as a primary endpoint and reported median survival for both the experimental and control arms, allowing calculation of the absolute difference between arms at approval. Trials that listed both OS and FS as primary endpoints were included in both endpoint categories.

Noticing in the summary shown in **Figures 5A and 5B** very similar hazard ratios for overall survival but clear differences between the FS hazard ratios – with a more granular look at lung cancer shown in **Figure 5C** – we explored in detail the results in the studies with mention of Progression-/Event-/Relapse-/Disease-free-survival [FS] and Overall Survival [OS] in approval letters. Often these were therapeutics in which the FS event outcome was clear in the approval letter, but the OS analysis was said to be immature or one in which the agency had determined “no trend towards a detriment was observed”. Note that the latter does not imply OS was at the time or eventually would be statistically favorable, since point estimates less than 1 that crossed the 1 boundary or even point estimates greater than 1 that crossed the 1 boundary could qualify for a “no trend towards a detriment was observed” designation. *With few exceptions, in those included in* ***Tables 1 and 2*** *we were able to find either in clintrials.gov, the literature or both information on OS and often final overall OS analyses and these are included in the tables.* Looking first at the FS analyses, the data in **Tables 1 and 2** align with the findings in **Figures 5A, 5B and 5C.** Compared to immunotherapies, the gains in the FS endpoints (months ΔFS) were longer and the point estimates and hazard ratios for the FS endpoints were statistically significantly lower with the non-immunotherapies in both first and second lines, consistent with the observations in **Figures 4A and 4C** that the FS outcomes recorded more favorable results with non-immunotherapies. We would also add that focusing on the HRs of the first line studies as shown in **Figures 5A**, the results achieved in the non-immunotherapy trials were statistically superior to those achieved in the immunotherapy studies as confirmed in **Figure 5B**. Again, here keep in mind the impact of redundant/me-too targeted therapy trials, potentially skewing results over the decade. Interestingly, with the non-immunotherapies, the HRs of studies conducted in first line were statistically indistinguishable from those in second line. However, as shown especially by the HRs in **Figures 5A and 5B**, the non-immunotherapy results were considerably less robust for the OS endpoint than the FS endpoints, with OS HRs statistically indistinguishable between non-immunotherapies and immunotherapies

**Table 1.**
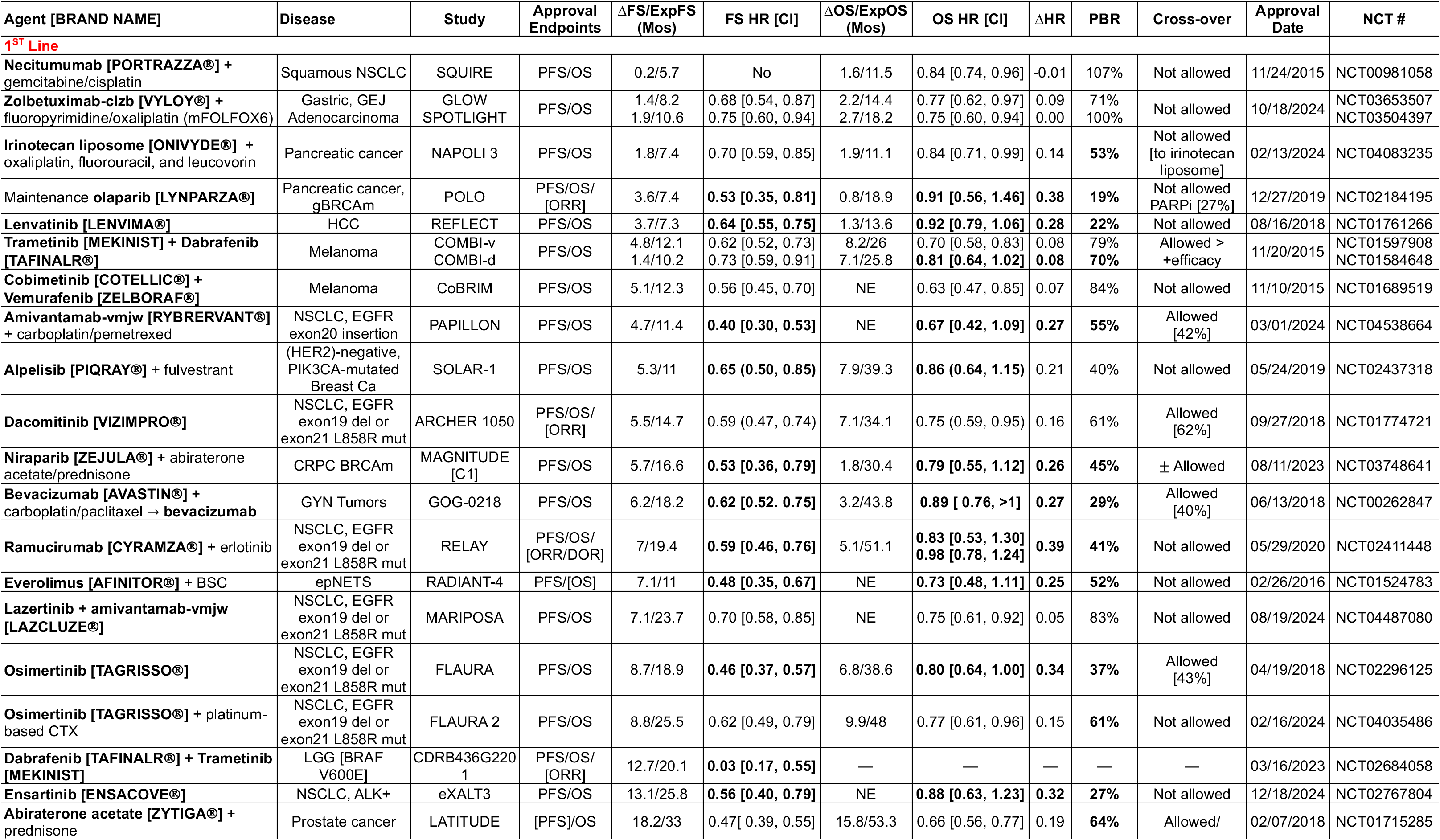
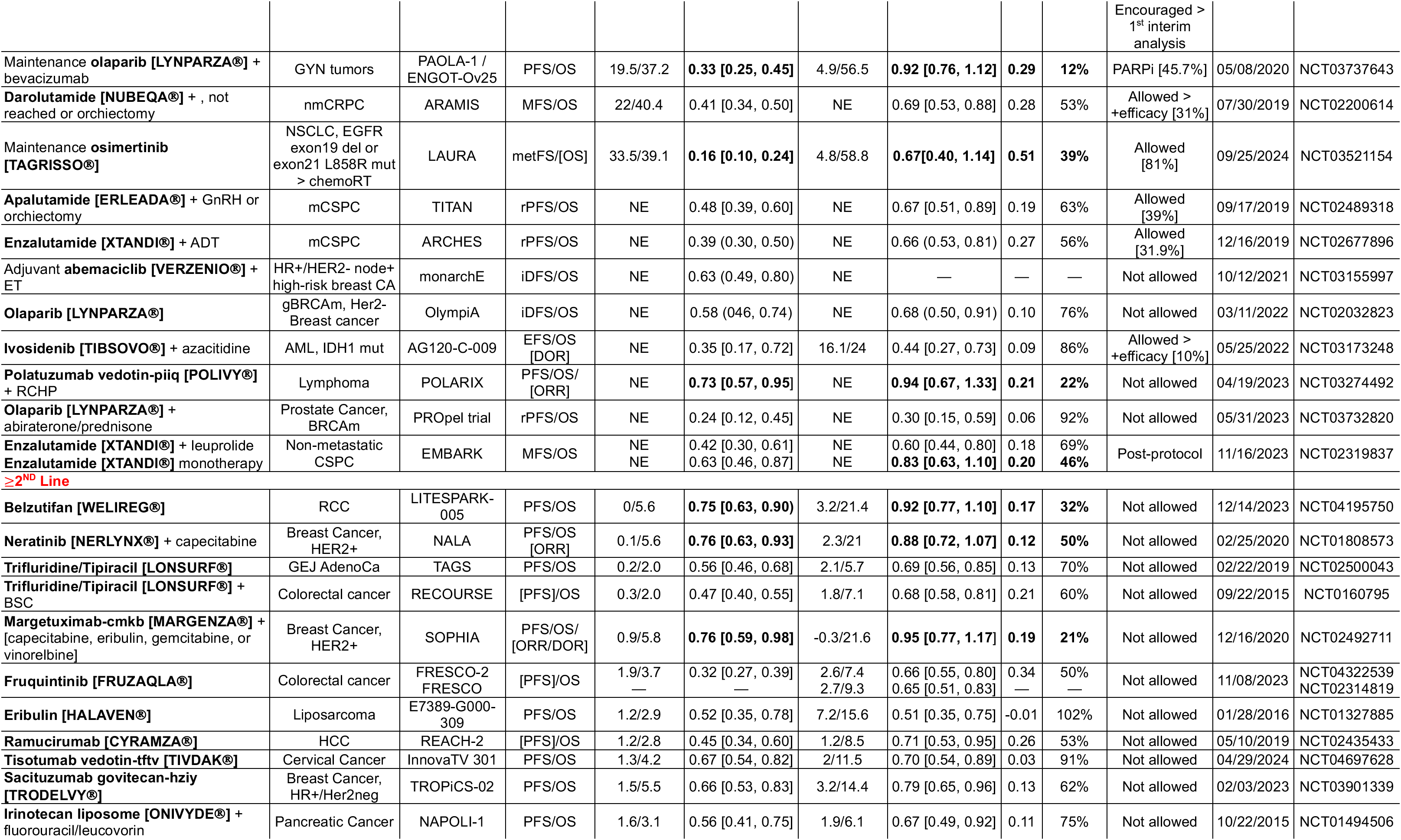
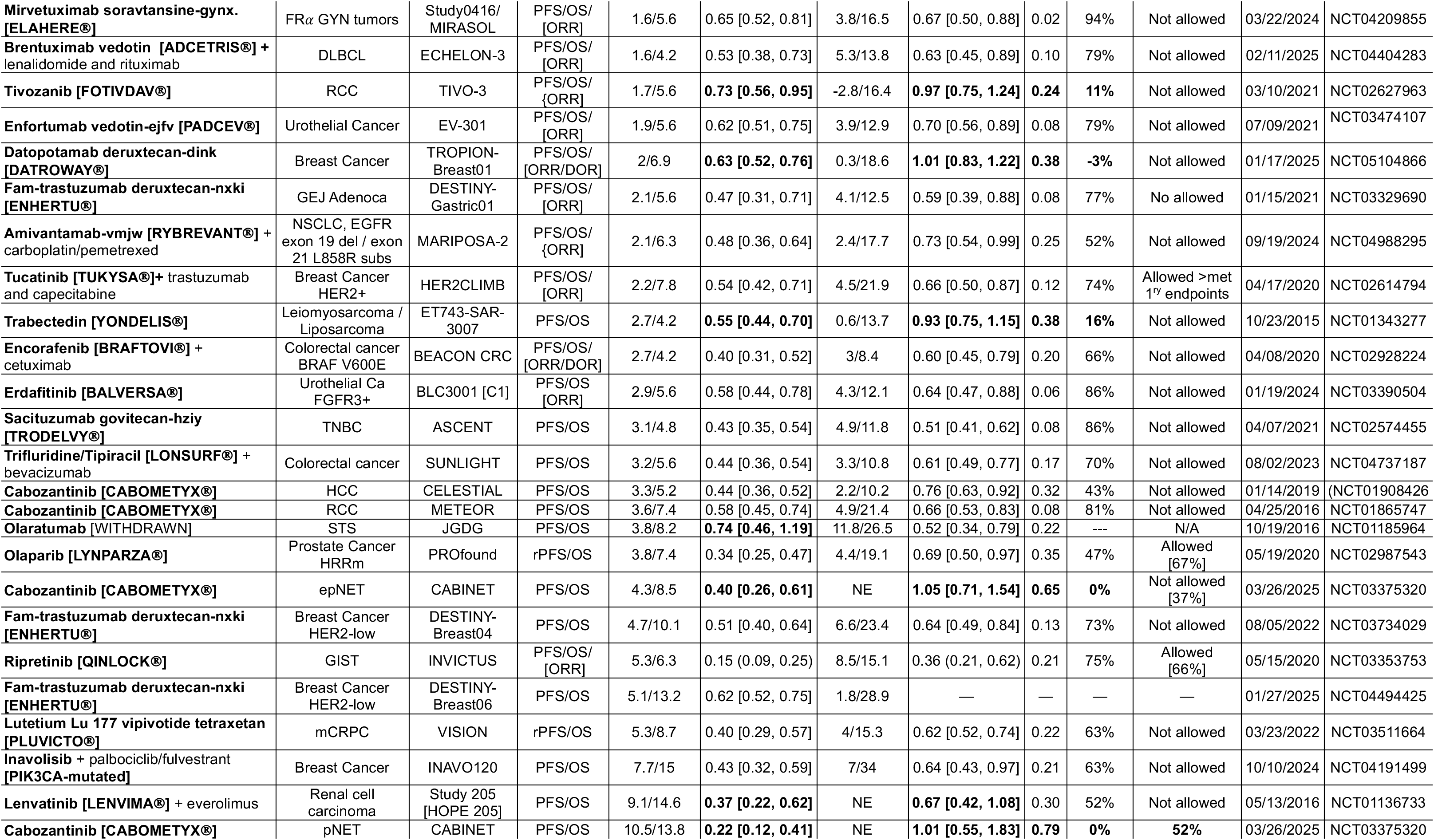
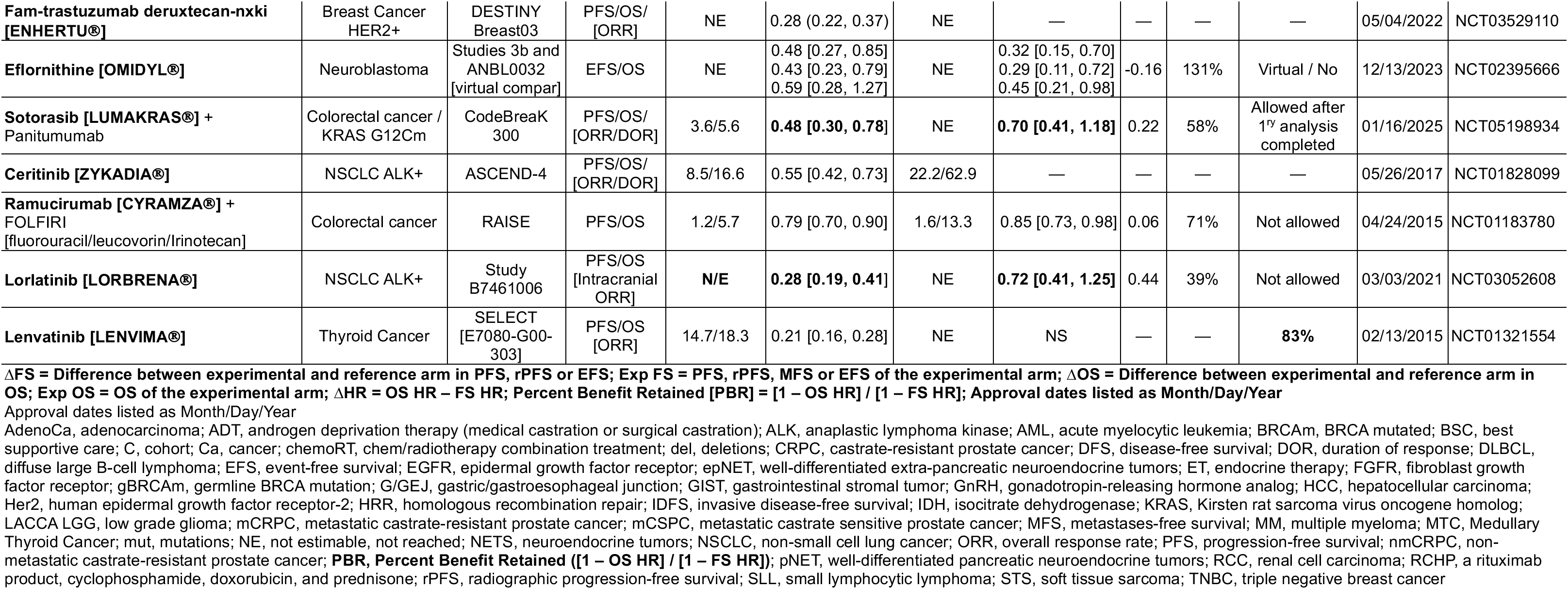
Non-Immunotherapy Approvals with Mention of Progression-/Event-/Relapse-/Disease-free-survival and Overall Survival in Approval Letter.

**Table 2.**
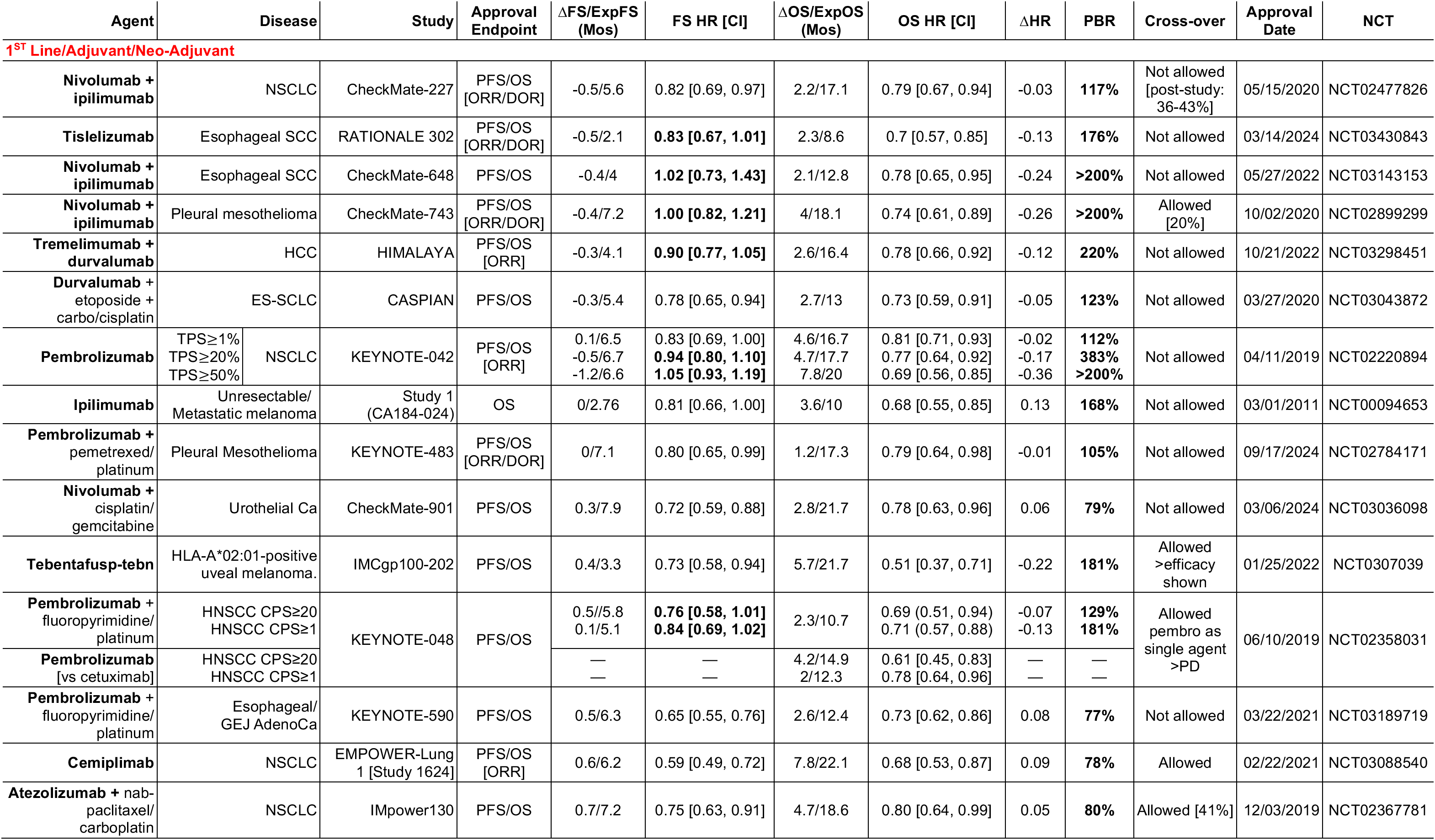
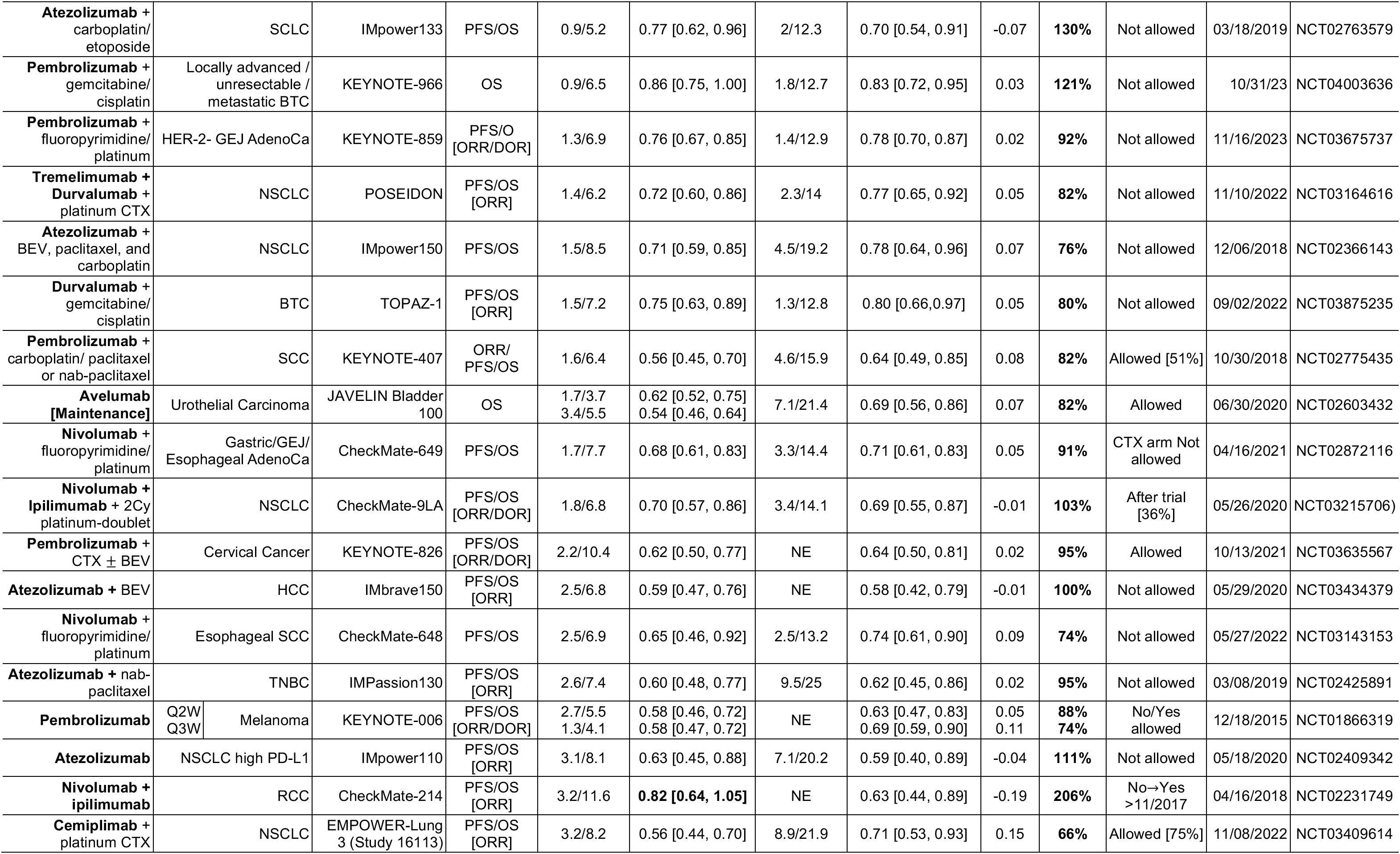
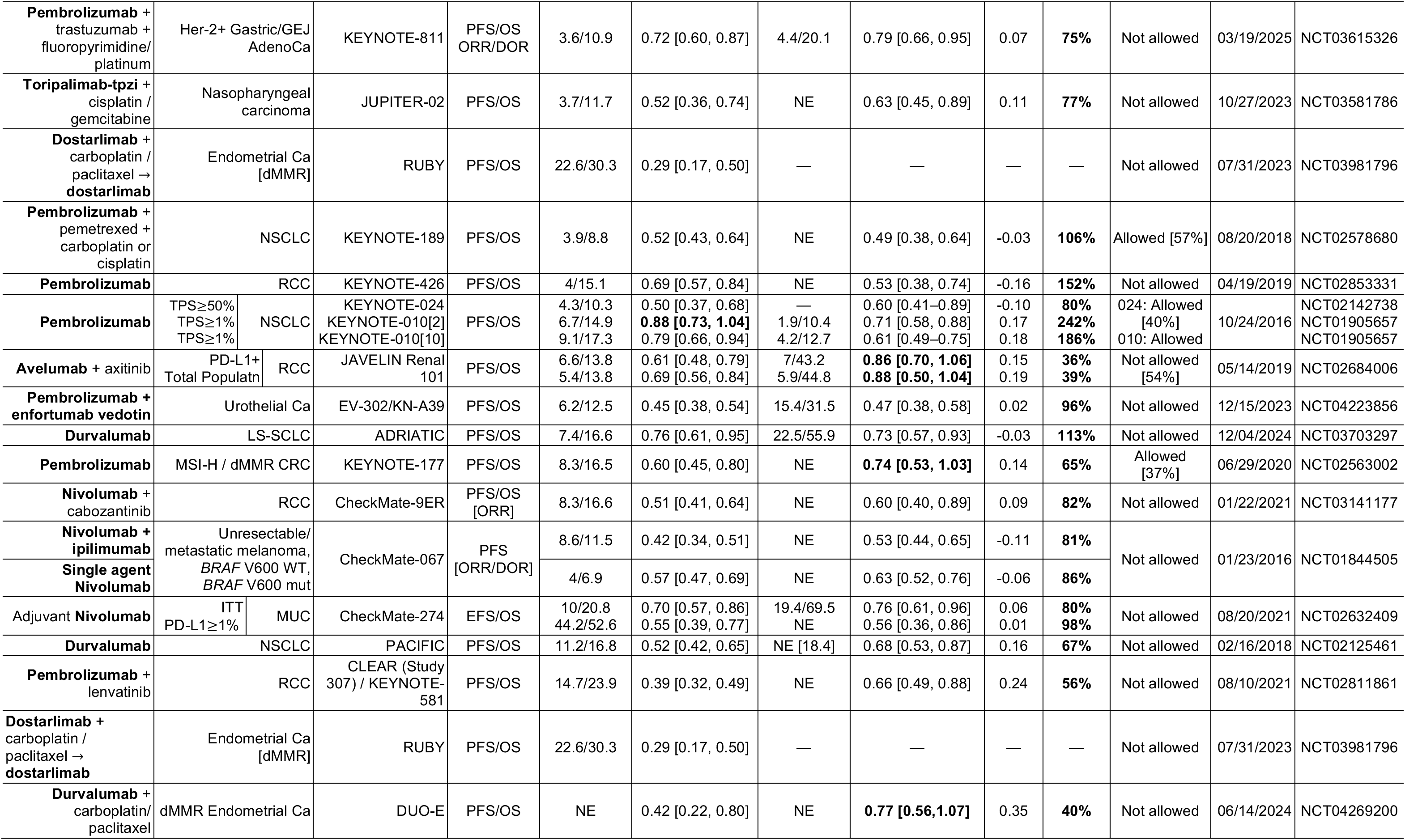
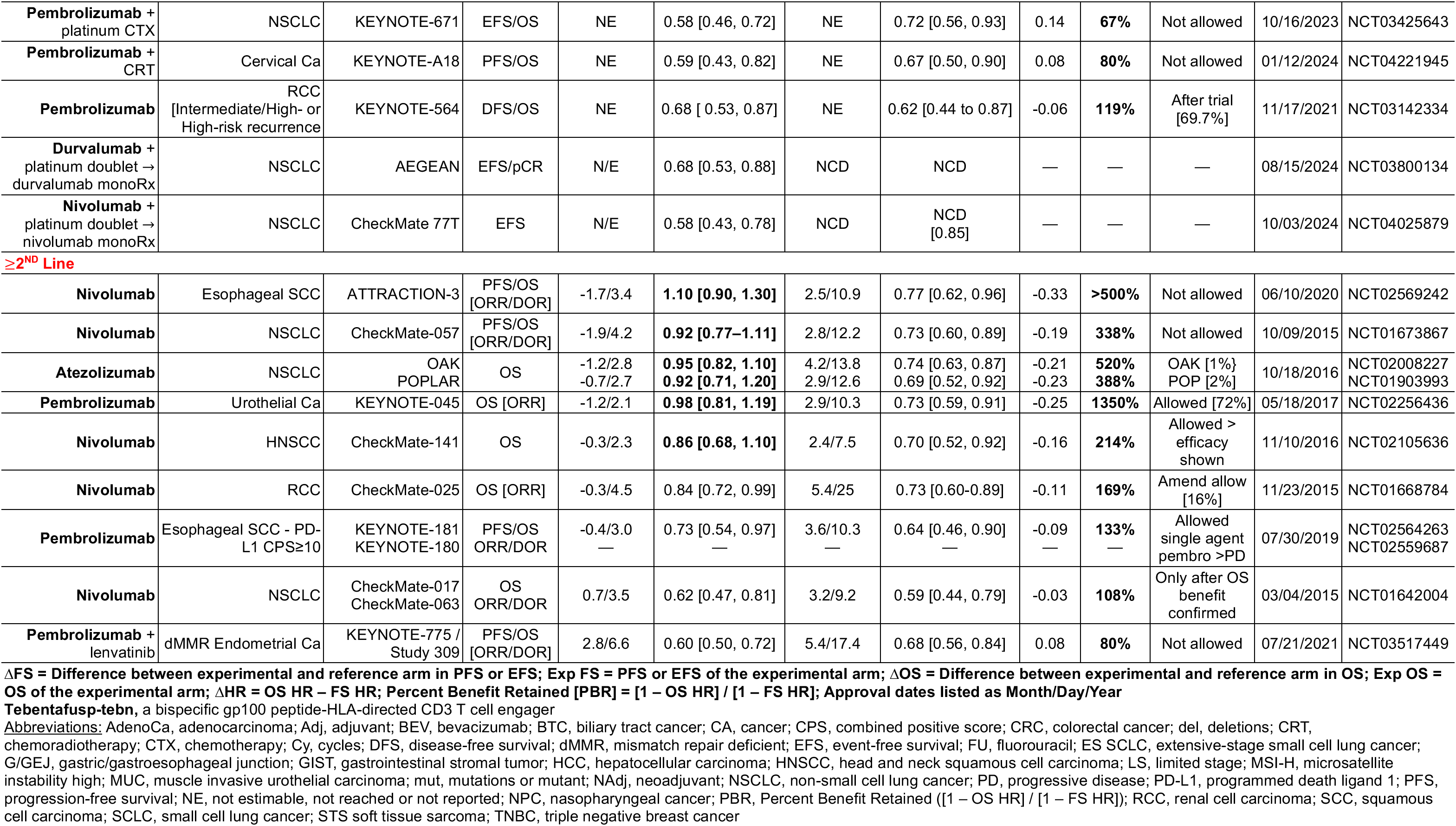
Immunotherapy Approvals with Mention of Progression-/Event-/Relapse-/Disease-free-survival and Overall Survival in Approval Letter.

| Agent | Disease |  | Study | Approval Endpoint | ΔFS/ExpFS (Mos) | FS HR [CI] | ΔOS/ExpOS (Mos) | OS HR [CI] | ΔHR | PBR | Cross-over | Approval Date | NCT |
| --- | --- | --- | --- | --- | --- | --- | --- | --- | --- | --- | --- | --- | --- |
| 1<sup>ST</sup> Line/Adjuvant/Neo-Adjuvant |  |  |  |  |  |  |  |  |  |  |  |  |  |
| Nivolumab + ipilimumab | NSCLC |  | CheckMate-227 | PFS/OS [ORR/DOR] | -0.5/5.6 | 0.82 [0.69, 0.97] | 2.2/17.1 | 0.79 [0.67, 0.94] | -0.03 | 117% | Not allowed [post-study: 36-43%] | 05/15/2020 | NCT02477826 |
| Tislelizumab | Esophageal SCC |  | RATIONALE 302 | PFS/OS [ORR/DOR] | -0.5/2.1 | 0.83 [0.67, 1.01] | 2.3/8.6 | 0.7 [0.57, 0.85] | -0.13 | 176% | Not allowed | 03/14/2024 | NCT03430843 |
| Nivolumab + ipilimumab | Esophageal SCC |  | CheckMate-648 | PFS/OS | -0.4/4 | 1.02 [0.73, 1.43] | 2.1/12.8 | 0.78 [0.65, 0.95] | -0.24 | >200% | Not allowed | 05/27/2022 | NCT03143153 |
| Nivolumab + ipilimumab | Pleural mesothelioma |  | CheckMate-743 | PFS/OS [ORR/DOR] | -0.4/7.2 | 1.00 [0.82, 1.21] | 4/18.1 | 0.74 [0.61, 0.89] | -0.26 | >200% | Allowed [20%] | 10/02/2020 | NCT02899299 |
| Tremelimumab + durvalumab | HCC |  | HIMALAYA | PFS/OS [ORR] | -0.3/4.1 | 0.90 [0.77, 1.05] | 2.6/16.4 | 0.78 [0.66, 0.92] | -0.12 | 220% | Not allowed | 10/21/2022 | NCT03298451 |
| Durvalumab + etoposide + carbo/cisplatin | ES-SCLC |  | CASPIAN | PFS/OS | -0.3/5.4 | 0.78 [0.65, 0.94] | 2.7/13 | 0.73 [0.59, 0.91] | -0.05 | 123% | Not allowed | 03/27/2020 | NCT03043872 |
| Pembrolizumab | TPS≥1%<br>TPS≥20%<br>TPS≥50% | NSCLC | KEYNOTE-042 | PFS/OS [ORR] | 0.1/6.5<br>-0.5/6.7<br>-1.2/6.6 | 0.83 [0.69, 1.00]<br>0.94 [0.80, 1.10]<br>1.05 [0.93, 1.19] | 4.6/16.7<br>4.7/17.7<br>7.8/20 | 0.81 [0.71, 0.93]<br>0.77 [0.64, 0.92]<br>0.69 [0.56, 0.85] | -0.02<br>-0.17<br>-0.36 | 112%<br>383%<br>>200% | Not allowed | 04/11/2019 | NCT02220894 |
| Ipilimumab | Unresectable/ Metastatic melanoma |  | Study 1 (CA184-024) | OS | 0/2.76 | 0.81 [0.66, 1.00] | 3.6/10 | 0.68 [0.55, 0.85] | 0.13 | 168% | Not allowed | 03/01/2011 | NCT00094653 |
| Pembrolizumab + pemetrexed/ platinum | Pleural Mesothelioma |  | KEYNOTE-483 | PFS/OS [ORR/DOR] | 0/7.1 | 0.80 [0.65, 0.99] | 1.2/17.3 | 0.79 [0.64, 0.98] | -0.01 | 105% | Not allowed | 09/17/2024 | NCT02784171 |
| Nivolumab + cisplatin/ gemcitabine | Urothelial Ca |  | CheckMate-901 | PFS/OS | 0.3/7.9 | 0.72 [0.59, 0.88] | 2.8/21.7 | 0.78 [0.63, 0.96] | 0.06 | 79% | Not allowed | 03/06/2024 | NCT03036098 |
| Tebentafusp-tebn | HLA-A*02:01-positive uveal melanoma. |  | IMCgp100-202 | PFS/OS | 0.4/3.3 | 0.73 [0.58, 0.94] | 5.7/21.7 | 0.51 [0.37, 0.71] | -0.22 | 181% | Allowed >efficacy shown | 01/25/2022 | NCT0307039 |
| Pembrolizumab + fluoropyrimidine/ platinum | HNSCC CPS≥20<br>HNSCC CPS≥1 |  | KEYNOTE-048 | PFS/OS | 0.5//5.8<br>0.1/5.1 | 0.76 [0.58, 1.01]<br>0.84 [0.69, 1.02] | 2.3/10.7 | 0.69 (0.51, 0.94)<br>0.71 (0.57, 0.88) | -0.07<br>-0.13 | 129%<br>181% | Allowed pembro as single agent >PD | 06/10/2019 | NCT02358031 |
| Pembrolizumab [vs cetuximab] | HNSCC CPS≥20<br>HNSCC CPS≥1 |  |  |  | —<br>— | —<br>— | 4.2/14.9<br>2/12.3 | 0.61 [0.45, 0.83]<br>0.78 [0.64, 0.96] | —<br>— | —<br>— |  |  |  |
| Pembrolizumab + fluoropyrimidine/ platinum | Esophageal/ GEJ AdenoCa |  | KEYNOTE-590 | PFS/OS | 0.5/6.3 | 0.65 [0.55, 0.76] | 2.6/12.4 | 0.73 [0.62, 0.86] | 0.08 | 77% | Not allowed | 03/22/2021 | NCT03189719 |
| Cemiplimab | NSCLC |  | EMPOWER-Lung 1 [Study 1624] | PFS/OS [ORR] | 0.6/6.2 | 0.59 [0.49, 0.72] | 7.8/22.1 | 0.68 [0.53, 0.87] | 0.09 | 78% | Allowed | 02/22/2021 | NCT03088540 |
| Atezolizumab + nab-paclitaxel/ carboplatin | NSCLC |  | IMpower130 | PFS/OS | 0.7/7.2 | 0.75 [0.63, 0.91] | 4.7/18.6 | 0.80 [0.64, 0.99] | 0.05 | 80% | Allowed [41%] | 12/03/2019 | NCT02367781 |
| **Atezolizumab** + carboplatin/etoposide | SCLC |  | IMpower133 | PFS/OS | 0.9/5.2 | 0.77 [0.62, 0.96] | 2/12.3 | 0.70 [0.54, 0.91] | -0.07 | **130%** | Not allowed | 03/18/2019 | NCT02763579 |
| **Pembrolizumab** + gemcitabine/cisplatin | Locally advanced / unresectable / metastatic BTC |  | KEYNOTE-966 | OS | 0.9/6.5 | 0.86 [0.75, 1.00] | 1.8/12.7 | 0.83 [0.72, 0.95] | 0.03 | **121%** | Not allowed | 10/31/23 | NCT04003636 |
| **Pembrolizumab** + fluoropyrimidine/platinum | HER-2- GEJ AdenoCa |  | KEYNOTE-859 | PFS/O [ORR/DOR] | 1.3/6.9 | 0.76 [0.67, 0.85] | 1.4/12.9 | 0.78 [0.70, 0.87] | 0.02 | **92%** | Not allowed | 11/16/2023 | NCT03675737 |
| **Tremelimumab + Durvalumab** + platinum CTX | NSCLC |  | POSEIDON | PFS/OS [ORR] | 1.4/6.2 | 0.72 [0.60, 0.86] | 2.3/14 | 0.77 [0.65, 0.92] | 0.05 | **82%** | Not allowed | 11/10/2022 | NCT03164616 |
| **Atezolizumab** + BEV, paclitaxel, and carboplatin | NSCLC |  | IMpower150 | PFS/OS | 1.5/8.5 | 0.71 [0.59, 0.85] | 4.5/19.2 | 0.78 [0.64, 0.96] | 0.07 | **76%** | Not allowed | 12/06/2018 | NCT02366143 |
| **Durvalumab** + gemcitabine/cisplatin | BTC |  | TOPAZ-1 | PFS/OS [ORR] | 1.5/7.2 | 0.75 [0.63, 0.89] | 1.3/12.8 | 0.80 [0.66, 0.97] | 0.05 | **80%** | Not allowed | 09/02/2022 | NCT03875235 |
| **Pembrolizumab** + carboplatin/ paclitaxel or nab-paclitaxel | SCC |  | KEYNOTE-407 | ORR/ PFS/OS | 1.6/6.4 | 0.56 [0.45, 0.70] | 4.6/15.9 | 0.64 [0.49, 0.85] | 0.08 | **82%** | Allowed [51%] | 10/30/2018 | NCT02775435 |
| **Avelumab [Maintenance]** | Urothelial Carcinoma |  | JAVELIN Bladder 100 | OS | 1.7/3.7<br>3.4/5.5 | 0.62 [0.52, 0.75]<br>0.54 [0.46, 0.64] | 7.1/21.4 | 0.69 [0.56, 0.86] | 0.07 | **82%** | Allowed | 06/30/2020 | NCT02603432 |
| **Nivolumab** + fluoropyrimidine/ platinum | Gastric/GEJ/ Esophageal AdenoCa |  | CheckMate-649 | PFS/OS | 1.7/7.7 | 0.68 [0.61, 0.83] | 3.3/14.4 | 0.71 [0.61, 0.83] | 0.05 | **91%** | CTX arm Not allowed | 04/16/2021 | NCT02872116 |
| **Nivolumab + Ipilimumab** + 2Cy platinum-doublet | NSCLC |  | CheckMate-9LA | PFS/OS [ORR/DOR] | 1.8/6.8 | 0.70 [0.57, 0.86] | 3.4/14.1 | 0.69 [0.55, 0.87] | -0.01 | **103%** | After trial [36%] | 05/26/2020 | NCT03215706) |
| **Pembrolizumab** + CTX ± BEV | Cervical Cancer |  | KEYNOTE-826 | PFS/OS [ORR/DOR] | 2.2/10.4 | 0.62 [0.50, 0.77] | NE | 0.64 [0.50, 0.81] | 0.02 | **95%** | Allowed | 10/13/2021 | NCT03635567 |
| **Atezolizumab** + BEV | HCC |  | IMbrave150 | PFS/OS [ORR] | 2.5/6.8 | 0.59 [0.47, 0.76] | NE | 0.58 [0.42, 0.79] | -0.01 | **100%** | Not allowed | 05/29/2020 | NCT03434379 |
| **Nivolumab** + fluoropyrimidine/ platinum | Esophageal SCC |  | CheckMate-648 | PFS/OS | 2.5/6.9 | 0.65 [0.46, 0.92] | 2.5/13.2 | 0.74 [0.61, 0.90] | 0.09 | **74%** | Not allowed | 05/27/2022 | NCT03143153 |
| **Atezolizumab** + nab-paclitaxel | TNBC |  | IMPassion130 | PFS/OS [ORR] | 2.6/7.4 | 0.60 [0.48, 0.77] | 9.5/25 | 0.62 [0.45, 0.86] | 0.02 | **95%** | Not allowed | 03/08/2019 | NCT02425891 |
| **Pembrolizumab** | Q2W/ Q3W | Melanoma | KEYNOTE-006 | PFS/OS [ORR/DOR] | 2.7/5.5<br>1.3/4.1 | 0.58 [0.46, 0.72]<br>0.58 [0.47, 0.72] | NE | 0.63 [0.47, 0.83]<br>0.69 [0.59, 0.90] | 0.05<br>0.11 | **88%**<br>**74%** | No/Yes allowed | 12/18/2015 | NCT01866319 |
| **Atezolizumab** | NSCLC high PD-L1 |  | IMpower110 | PFS/OS [ORR] | 3.1/8.1 | 0.63 [0.45, 0.88] | 7.1/20.2 | 0.59 [0.40, 0.89] | -0.04 | **111%** | Not allowed | 05/18/2020 | NCT02409342 |
| **Nivolumab + ipilimumab** | RCC |  | CheckMate-214 | PFS/OS [ORR] | 3.2/11.6 | **0.82 [0.64, 1.05]** | NE | 0.63 [0.44, 0.89] | -0.19 | **206%** | No→Yes >11/2017 | 04/16/2018 | NCT02231749 |
| **Cemiplimab** + platinum CTX | NSCLC |  | EMPOWER-Lung 3 (Study 16113) | PFS/OS [ORR] | 3.2/8.2 | 0.56 [0.44, 0.70] | 8.9/21.9 | 0.71 [0.53, 0.93] | 0.15 | **66%** | Allowed [75%] | 11/08/2022 | NCT03409614 |
| **Pembrolizumab + trastuzumab + fluoropyrimidine/ platinum** | Her-2+ Gastric/GEJ AdenoCa |  | KEYNOTE-811 | PFS/OS<br>ORR/DOR | 3.6/10.9 | 0.72 [0.60, 0.87] | 4.4/20.1 | 0.79 [0.66, 0.95] | 0.07 | **75%** | Not allowed | 03/19/2025 | NCT03615326 |
| **Toripalimab-tpzi + cisplatin / gemcitabine** | Nasopharyngeal carcinoma |  | JUPITER-02 | PFS/OS | 3.7/11.7 | 0.52 [0.36, 0.74] | NE | 0.63 [0.45, 0.89] | 0.11 | **77%** | Not allowed | 10/27/2023 | NCT03581786 |
| **Dostarlimab + carboplatin / paclitaxel → dostarlimab** | Endometrial Ca [dMMR] |  | RUBY | PFS/OS | 22.6/30.3 | 0.29 [0.17, 0.50] | — | — | — | — | Not allowed | 07/31/2023 | NCT03981796 |
| **Pembrolizumab + pemetrexed + carboplatin or cisplatin** | NSCLC |  | KEYNOTE-189 | PFS/OS | 3.9/8.8 | 0.52 [0.43, 0.64] | NE | 0.49 [0.38, 0.64] | -0.03 | **106%** | Allowed [57%] | 08/20/2018 | NCT02578680 |
| **Pembrolizumab** | RCC |  | KEYNOTE-426 | PFS/OS | 4/15.1 | 0.69 [0.57, 0.84] | NE | 0.53 [0.38, 0.74] | -0.16 | **152%** | Not allowed | 04/19/2019 | NCT02853331 |
| **Pembrolizumab** | TPS≥50% | NSCLC | KEYNOTE-024 | PFS/OS | 4.3/10.3 | 0.50 [0.37, 0.68] | — | 0.60 [0.41–0.89] | -0.10 | **80%** | 024: Allowed [40%]<br>010: Allowed | 10/24/2016 | NCT02142738<br>NCT01905657<br>NCT01905657 |
|  | TPS≥1% |  | KEYNOTE-010[2] |  | 6.7/14.9 | **0.88 [0.73, 1.04]** | 1.9/10.4 | 0.71 [0.58, 0.88] | 0.17 | **242%** |  |  |  |
|  | TPS≥1% |  | KEYNOTE-010[10] |  | 9.1/17.3 | 0.79 [0.66, 0.94] | 4.2/12.7 | 0.61 [0.49–0.75] | 0.18 | **186%** |  |  |  |
| **Avelumab + axitinib** | PD-L1+<br>Total Populatn | RCC | JAVELIN Renal 101 | PFS/OS | 6.6/13.8<br>5.4/13.8 | 0.61 [0.48, 0.79]<br>0.69 [0.56, 0.84] | 7/43.2<br>5.9/44.8 | **0.86 [0.70, 1.06]**<br>**0.88 [0.50, 1.04]** | 0.15<br>0.19 | **36%**<br>**39%** | Not allowed [54%] | 05/14/2019 | NCT02684006 |
| **Pembrolizumab + enfortumab vedotin** | Urothelial Ca |  | EV-302/KN-A39 | PFS/OS | 6.2/12.5 | 0.45 [0.38, 0.54] | 15.4/31.5 | 0.47 [0.38, 0.58] | 0.02 | **96%** | Not allowed | 12/15/2023 | NCT04223856 |
| **Durvalumab** | LS-SCLC |  | ADRIATIC | PFS/OS | 7.4/16.6 | 0.76 [0.61, 0.95] | 22.5/55.9 | 0.73 [0.57, 0.93] | -0.03 | **113%** | Not allowed | 12/04/2024 | NCT03703297 |
| **Pembrolizumab** | MSI-H / dMMR CRC |  | KEYNOTE-177 | PFS/OS | 8.3/16.5 | 0.60 [0.45, 0.80] | NE | **0.74 [0.53, 1.03]** | 0.14 | **65%** | Allowed [37%] | 06/29/2020 | NCT02563002 |
| **Nivolumab + cabozantinib** | RCC |  | CheckMate-9ER | PFS/OS [ORR] | 8.3/16.6 | 0.51 [0.41, 0.64] | NE | 0.60 [0.40, 0.89] | 0.09 | **82%** | Not allowed | 01/22/2021 | NCT03141177 |
| **Nivolumab + ipilimumab** | Unresectable/ metastatic melanoma, BRAF V600 WT, BRAF V600 mut |  | CheckMate-067 | PFS [ORR/DOR] | 8.6/11.5 | 0.42 [0.34, 0.51] | NE | 0.53 [0.44, 0.65] | -0.11 | **81%** | Not allowed | 01/23/2016 | NCT01844505 |
| **Single agent Nivolumab** |  |  |  |  | 4/6.9 | 0.57 [0.47, 0.69] | NE | 0.63 [0.52, 0.76] | -0.06 | **86%** |  |  |  |
| Adjuvant **Nivolumab** | ITT<br>PD-L1≥1% | MUC | CheckMate-274 | EFS/OS | 10/20.8<br>44.2/52.6 | 0.70 [0.57, 0.86]<br>0.55 [0.39, 0.77] | 19.4/69.5<br>NE | 0.76 [0.61, 0.96]<br>0.56 [0.36, 0.86] | 0.06<br>0.01 | **80%**<br>**98%** | Not allowed | 08/20/2021 | NCT02632409 |
| **Durvalumab** | NSCLC |  | PACIFIC | PFS/OS | 11.2/16.8 | 0.52 [0.42, 0.65] | NE [18.4] | 0.68 [0.53, 0.87] | 0.16 | **67%** | Not allowed | 02/16/2018 | NCT02125461 |
| **Pembrolizumab + lenvatinib** | RCC |  | CLEAR (Study 307) / KEYNOTE-581 | PFS/OS | 14.7/23.9 | 0.39 [0.32, 0.49] | NE | 0.66 [0.49, 0.88] | 0.24 | **56%** | Not allowed | 08/10/2021 | NCT02811861 |
| **Dostarlimab + carboplatin / paclitaxel → dostarlimab** | Endometrial Ca [dMMR] |  | RUBY | PFS/OS | 22.6/30.3 | 0.29 [0.17, 0.50] | — | — | — | — | Not allowed | 07/31/2023 | NCT03981796 |
| **Durvalumab + carboplatin/ paclitaxel** | dMMR Endometrial Ca |  | DUO-E | PFS/OS | NE | 0.42 [0.22, 0.80] | NE | **0.77 [0.56,1.07]** | 0.35 | **40%** | Not allowed | 06/14/2024 | NCT04269200 |

|  |  |  |  |  |  |  |  |  |  |  |  |  |
| --- | --- | --- | --- | --- | --- | --- | --- | --- | --- | --- | --- | --- |
| **Pembrolizumab + platinum CTX** | NSCLC | KEYNOTE-671 | EFS/OS | NE | 0.58 [0.46, 0.72] | NE | 0.72 [0.56, 0.93] | 0.14 | **67%** | Not allowed | 10/16/2023 | NCT03425643 |
| **Pembrolizumab + CRT** | Cervical Ca | KEYNOTE-A18 | PFS/OS | NE | 0.59 [0.43, 0.82] | NE | 0.67 [0.50, 0.90] | 0.08 | **80%** | Not allowed | 01/12/2024 | NCT04221945 |
| **Pembrolizumab** | RCC<br>[Intermediate/High- or High-risk recurrence] | KEYNOTE-564 | DFS/OS | NE | 0.68 [0.53, 0.87] | NE | 0.62 [0.44 to 0.87] | -0.06 | **119%** | After trial [69.7%] | 11/17/2021 | NCT03142334 |
| **Durvalumab + platinum doublet → durvalumab monoRx** | NSCLC | AEGEAN | EFS/pCR | N/E | 0.68 [0.53, 0.88] | NCD | NCD | — | — | — | 08/15/2024 | NCT03800134 |
| **Nivolumab + platinum doublet → nivolumab monoRx** | NSCLC | CheckMate 77T | EFS | N/E | 0.58 [0.43, 0.78] | NCD | NCD [0.85] | — | — | — | 10/03/2024 | NCT04025879 |

≥2<sup>ND</sup> Line
|  |  |  |  |  |  |  |  |  |  |  |  |  |
| --- | --- | --- | --- | --- | --- | --- | --- | --- | --- | --- | --- | --- |
| **Nivolumab** | Esophageal SCC | ATTRACTION-3 | PFS/OS [ORR/DOR] | -1.7/3.4 | **1.10 [0.90, 1.30]** | 2.5/10.9 | 0.77 [0.62, 0.96] | -0.33 | **>500%** | Not allowed | 06/10/2020 | NCT02569242 |
| **Nivolumab** | NSCLC | CheckMate-057 | PFS/OS [ORR/DOR] | -1.9/4.2 | **0.92 [0.77–1.11]** | 2.8/12.2 | 0.73 [0.60, 0.89] | -0.19 | **338%** | Not allowed | 10/09/2015 | NCT01673867 |
| **Atezolizumab** | NSCLC | OAK<br>POPLAR | OS | -1.2/2.8<br>-0.7/2.7 | **0.95 [0.82, 1.10]**<br>**0.92 [0.71, 1.20]** | 4.2/13.8<br>2.9/12.6 | 0.74 [0.63, 0.87]<br>0.69 [0.52, 0.92] | -0.21<br>-0.23 | **520%**<br>**388%** | OAK [1%]<br>POP [2%] | 10/18/2016 | NCT02008227<br>NCT01903993 |
| **Pembrolizumab** | Urothelial Ca | KEYNOTE-045 | OS [ORR] | -1.2/2.1 | **0.98 [0.81, 1.19]** | 2.9/10.3 | 0.73 [0.59, 0.91] | -0.25 | **1350%** | Allowed [72%] | 05/18/2017 | NCT02256436 |
| **Nivolumab** | HNSCC | CheckMate-141 | OS | -0.3/2.3 | **0.86 [0.68, 1.10]** | 2.4/7.5 | 0.70 [0.52, 0.92] | -0.16 | **214%** | Allowed > efficacy shown | 11/10/2016 | NCT02105636 |
| **Nivolumab** | RCC | CheckMate-025 | OS [ORR] | -0.3/4.5 | 0.84 [0.72, 0.99] | 5.4/25 | 0.73 [0.60-0.89] | -0.11 | **169%** | Amend allow [16%] | 11/23/2015 | NCT01668784 |
| **Pembrolizumab** | Esophageal SCC - PD-L1 CPS≥10 | KEYNOTE-181<br>KEYNOTE-180 | PFS/OS<br>ORR/DOR | -0.4/3.0<br>— | 0.73 [0.54, 0.97]<br>— | 3.6/10.3<br>— | 0.64 [0.46, 0.90]<br>— | -0.09<br>— | **133%**<br>— | Allowed single agent pembro >PD | 07/30/2019 | NCT02564263<br>NCT02559687 |
| **Nivolumab** | NSCLC | CheckMate-017<br>CheckMate-063 | OS<br>ORR/DOR | 0.7/3.5 | 0.62 [0.47, 0.81] | 3.2/9.2 | 0.59 [0.44, 0.79] | -0.03 | **108%** | Only after OS benefit confirmed | 03/04/2015 | NCT01642004 |
| **Pembrolizumab + lenvatinib** | dMMR Endometrial Ca | KEYNOTE-775 /<br>Study 309 | PFS/OS [ORR/DOR] | 2.8/6.6 | 0.60 [0.50, 0.72] | 5.4/17.4 | 0.68 [0.56, 0.84] | 0.08 | **80%** | Not allowed | 07/21/2021 | NCT03517449 |

ΔFS = Difference between experimental and reference arm in PFS or EFS; Exp FS = PFS or EFS of the experimental arm; ΔOS = Difference between experimental and reference arm in OS; Exp OS = OS of the experimental arm; ΔHR = OS HR – FS HR; Percent Benefit Retained [PBR] = [1 – OS HR] / [1 – FS HR]; Approval dates listed as Month/Day/Year
**Tebentafusp-tebn**, a bispecific gp100 peptide-HLA-directed CD3 T cell engager
**Abbreviations:** AdenoCa, adenocarcinoma; Adj, adjuvant; BEV, bevacizumab; BTC, biliary tract cancer; CA, cancer; CPS, combined positive score; CRC, colorectal cancer; del, deletions; CRT, chemoradiotherapy; CTX, chemotherapy; Cy, cycles; DFS, disease-free survival; dMMR, mismatch repair deficient; EFS, event-free survival; FU, fluorouracil; ES SCLC, extensive-stage small cell lung cancer; G/GEJ, gastric/gastroesophageal junction; GIST, gastrointestinal stromal tumor; HCC, hepatocellular carcinoma; HNSCC, head and neck squamous cell carcinoma; LS, limited stage; MSI-H, microsatellite instability high; MUC, muscle invasive urothelial carcinoma; mut, mutations or mutant; NAdj, neoadjuvant; NSCLC, non-small cell lung cancer; PD, progressive disease; PD-L1, programmed death ligand 1; PFS, progression-free survival; NE, not estimable, not reached or not reported; NPC, nasopharyngeal cancer; PBR, Percent Benefit Retained ([1 – OS HR] / [1 – FS HR]); RCC, renal cell carcinoma; SCC, squamous cell carcinoma; SCLC, small cell lung cancer; STS soft tissue sarcoma; TNBC, triple negative breast cancer

Given these results we dove deeper into the data looking for possible explanations. Recognizing that a statistically significant FS advantage in the non-immunotherapy trials had translated poorly into overall survival, we looked to the individual trials to see if all or an identifiable fraction had faltered. Investigators frequently describe the benefits of a particular therapy by touting its ability to reduce the risk of an adverse event by a given percentage. For example, a therapy with a HR for PFS of 0.72 is said to have successfully reduced the risk of progression or death by 28% [(1 – 0.72) X 100); or an intervention with an OS HR of 0.79 is reported to have reduced the risk of death by 21% [(1 – 0.79) X 100] [although actually they reduce the rate at which the risks occur not the risks overall]. Pivoting from this frequently used description we calculated the percent of benefit claimed by the FS HR that had translated to the OS HR, describing it as the precent of benefit translated (PBT). For each study, we calculated the fraction of the FS benefit that had translated to overall survival (OS) benefit as [1 – HROS / 1 – HRPFS] X100 with hazard ratios (HRs) taken from the approval letter, clintrials.gov or relevant publications [Note these results were remarkably similar to (log [HROS] / log [HRPFS]) X 100 that might be said to be more correct but we chose our approach as one more intuitive to clinicians]. Assessing the non-immunotherapy trials, the median fraction translated was 61%, an attenuation not observed with immunotherapies [**Figure 6A**] where most of the FS benefit translated to the OS outcome. As shown in **Figure 6B**, this difference in outcome between non-immunotherapies and immunotherapies was highly statistically different and was seen with therapies in both first and second line [**Figure 6C**] and with immunotherapies across diseases [**Figure 6D**].

**Figure 6.**
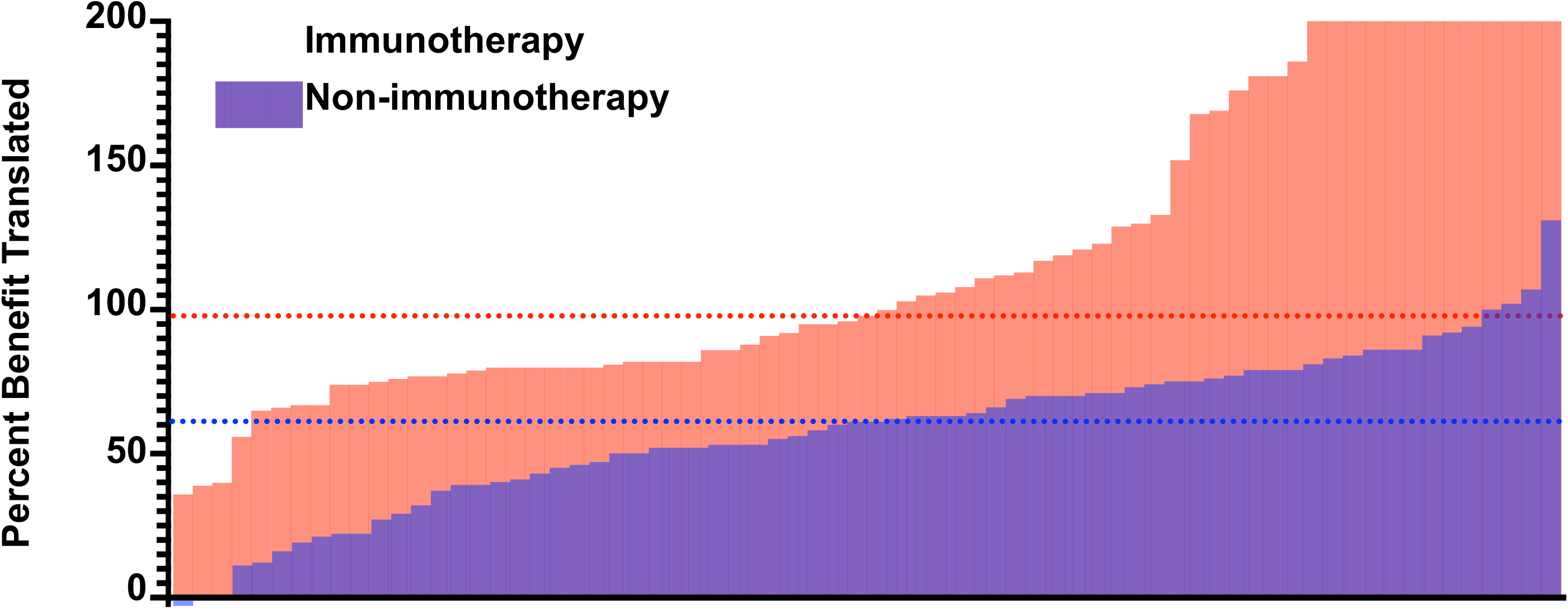
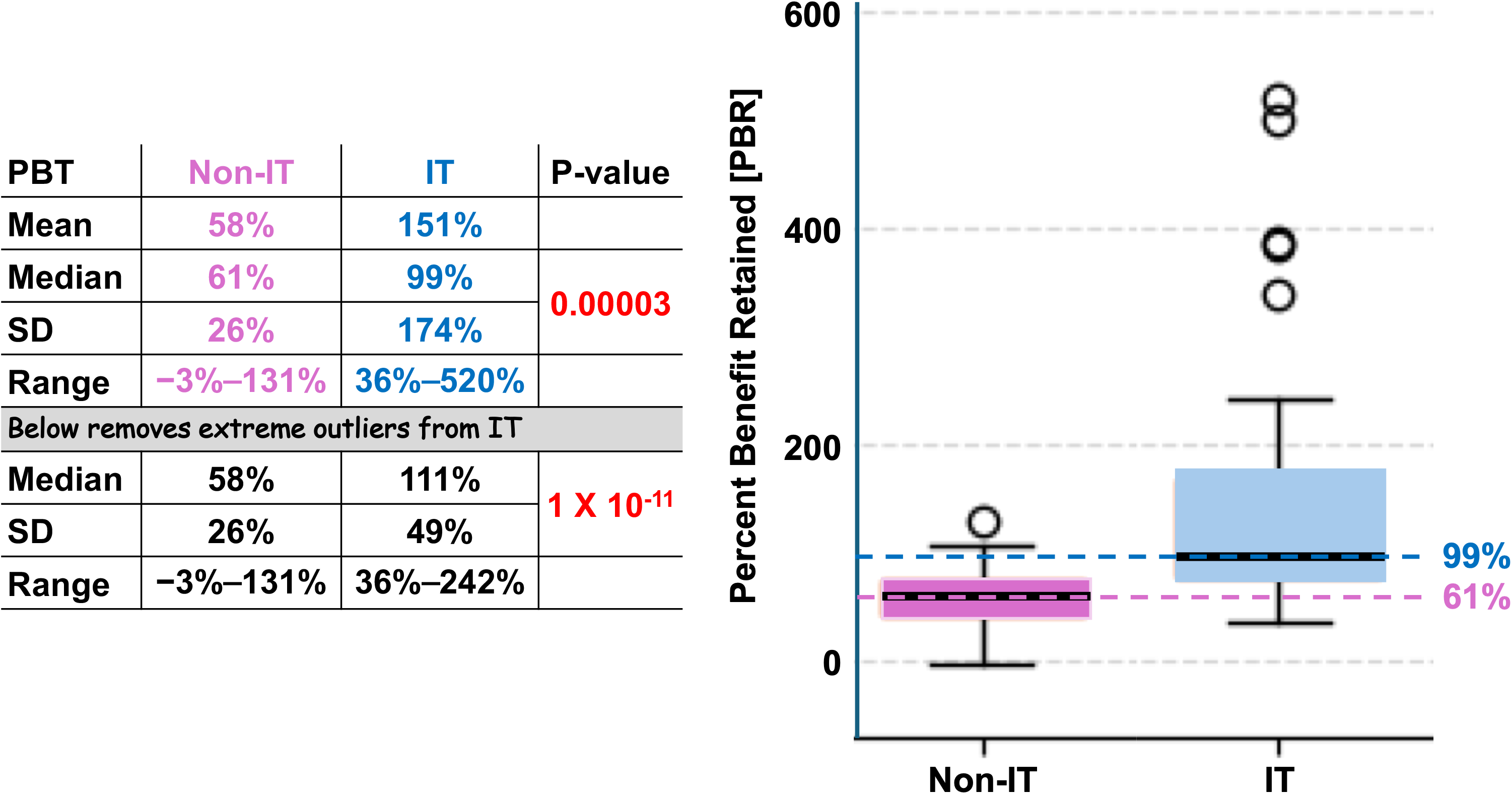
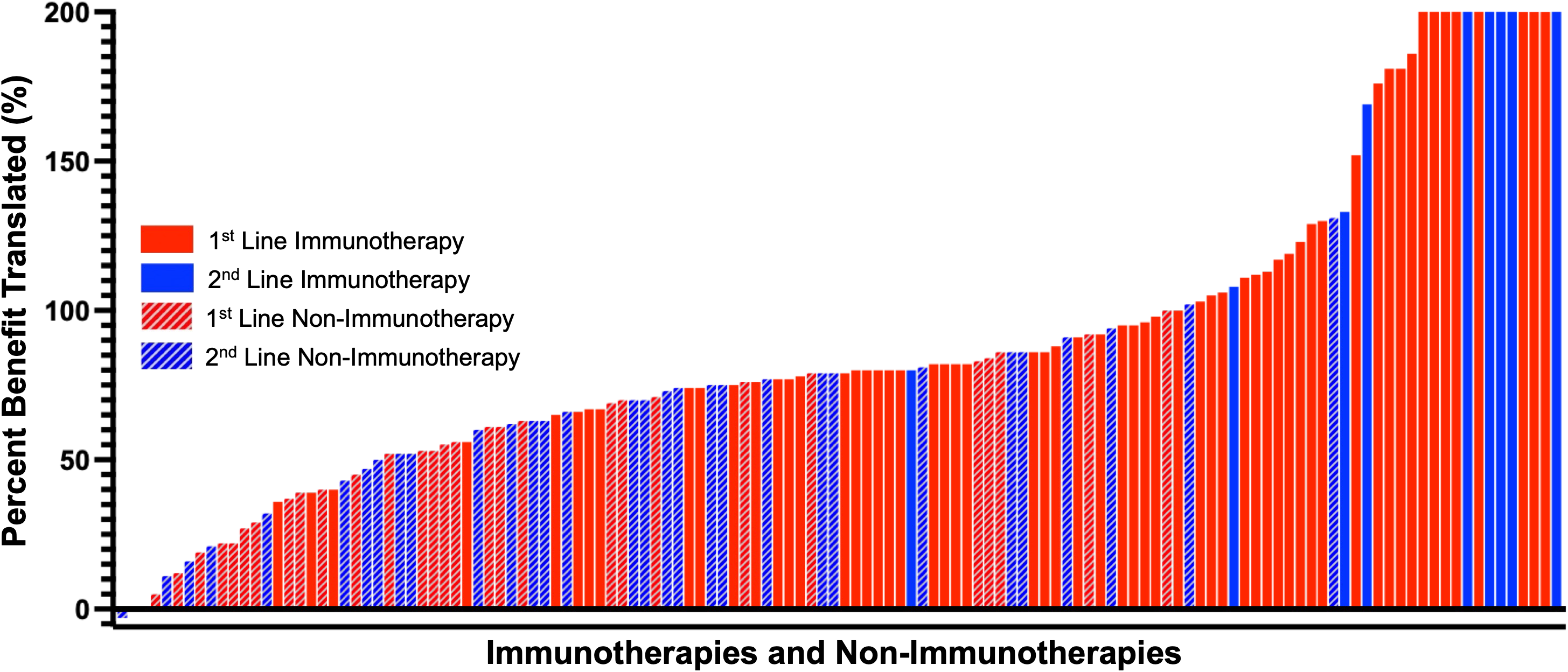
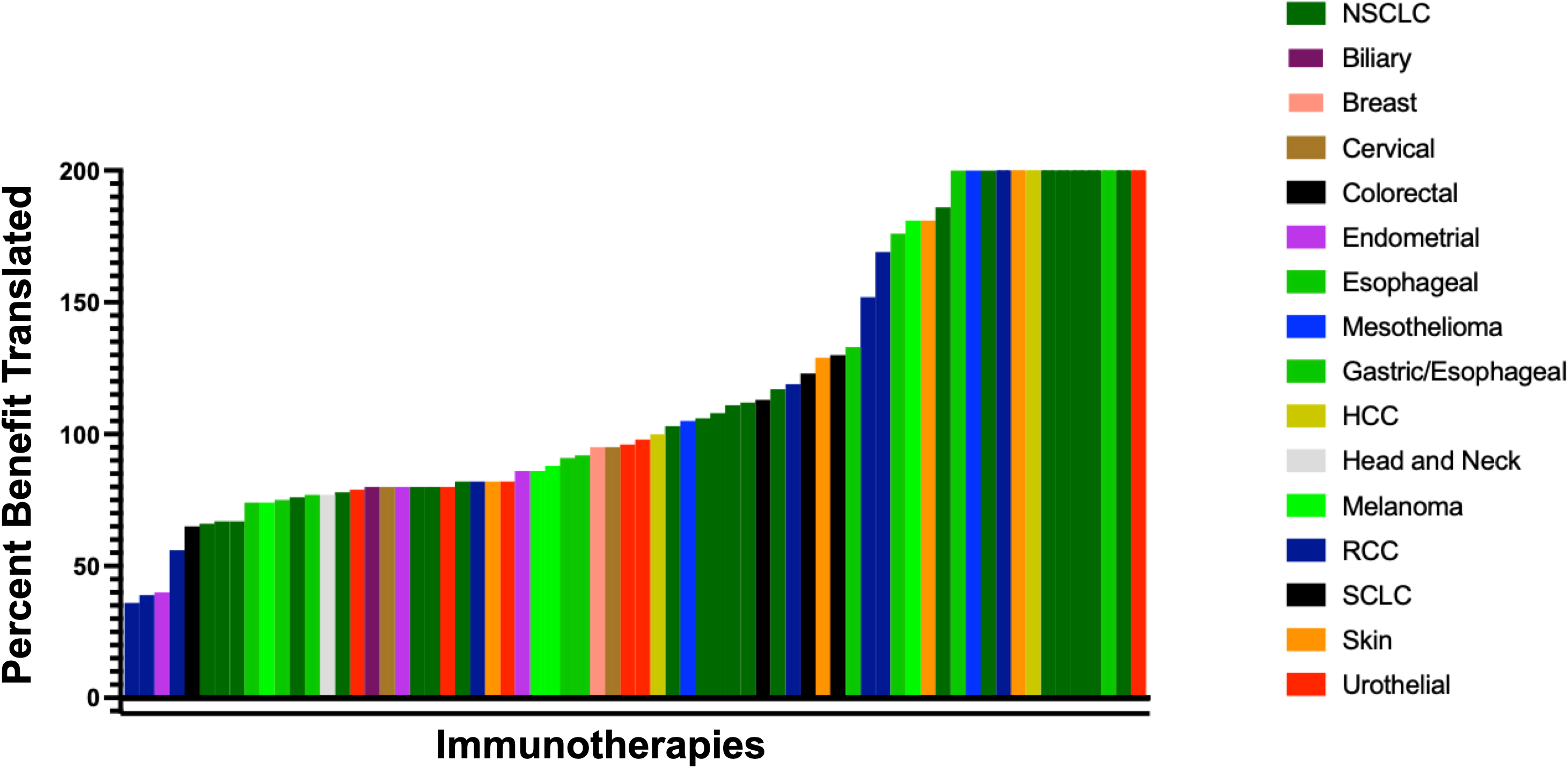
Immunotherapies translate a greater precent of their PFS gains to OS. [**A**] Percent of benefit in the FS endpoint translated [PBT] to the OS endpoint is seen to be markedly greater and [**B**] statistically superior for immunotherapies than for non-immunotherapies. [**C**] This translation of benefit is shown to be better for immunotherapies and non-immunotherapies independent of line of therapy and [**D**] independent of disease being treated. Note: For each study, the proportion of the FS benefit translated to benefit in overall survival (OS) was calculated as [1 – HROS / 1 – HRPFS] X100 with hazard ratios (HRs) taken from the approval letter, clintrials.gov or relevant publications [Note these results were remarkably similar to (log [HROS] / log [HRPFS]) X 100 that might be said to be more correct but we chose our approach as one more intuitive to clinicians]. PBT was calculated for each trial that listed both PFS and OS as primary endpoints and reported final median survival values for both the experimental and control arms for both endpoints.

Amongst the 78 non-immunotherapy trials summarized in **Table 1**, in 26 (33.3%) the experimental therapy did not achieve a statistically significant OS advantage over the reference, a failure rate much worse than that of the immunotherapy studies where only three [4%] of the approved studies with mention of PFS and OS in the approval letter did not achieve a statistically significant OS difference. Crossover, commonly invoked as an explanation for the disappearance of an advantage moving from the FS endpoint to the OS is an unlikely explanation. While crossover was allowed in 17 of the 74 non-immunotherapy studies (23%), it was allowed in only 10 of the 26 studies without a significant OS advantage and in only 6 of these was there crossover of 40% or higher. As shown in **Figure 7A**, unsurprisingly, non-immunotherapy trials achieving statistically significant OS benefits had substantially larger treatment effects than therapies in trials that did not achieve a statistically significant OS improvement, with median OS hazard ratios of 0.67 versus 0.88 (P <10⁻⁶). Note that despite some overlap in fact the two populations – those with or without a statistically significant OS outcome – do not appear as two ends of a continuum but rather two distinct populations with different outcomes.

**Figure 7.**
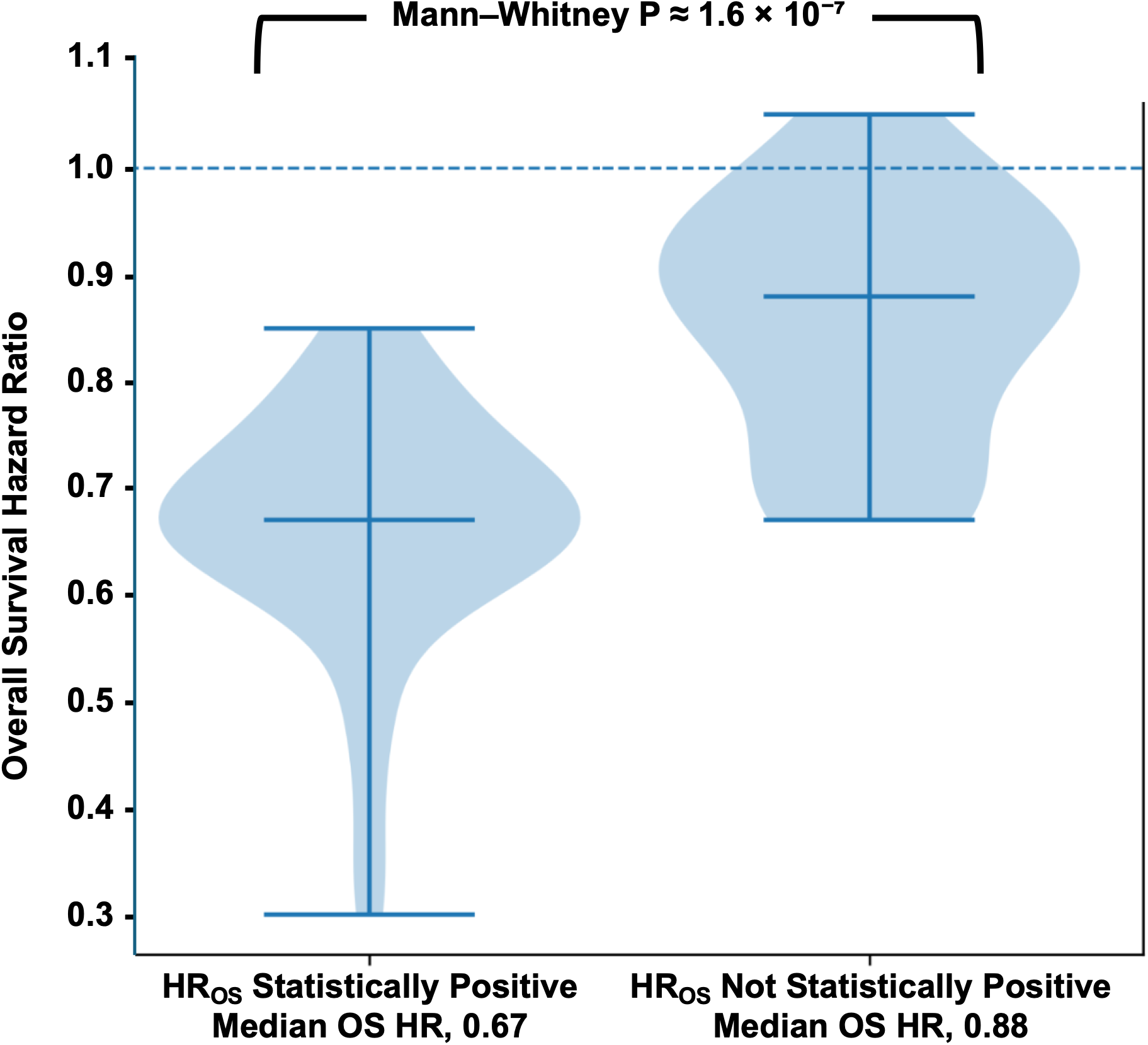
Amongst the non-immunotherapy trials, therapies achieving statistically significant OS benefits had substantially larger treatment effects than therapies in trials that did not achieve a statistically significant OS improvement. Despite some overlap in fact, they emerge as two different populations rather than a continuum.

Finally, although to date no trial has compared two checkpoint inhibitors, much less conducted a direct comparison of an anti-PD-1 to an anti-PD-L1 agent, we compared in an exploratory manner, outcomes with anti-PD-1 versus anti-PD-L1 agents again in studies where FS and OS endpoints had been referred to in the FDA approval letter. Recognizing this comparison can be criticized as less-than-ideal but keenly aware this will never be done prospectively, as shown in **Figure 8A** the cancers addressed have been similar and as shown in **Figure 8B** have achieved comparable effectiveness as measured by comparable hazard ratios.

**Figure 8.**
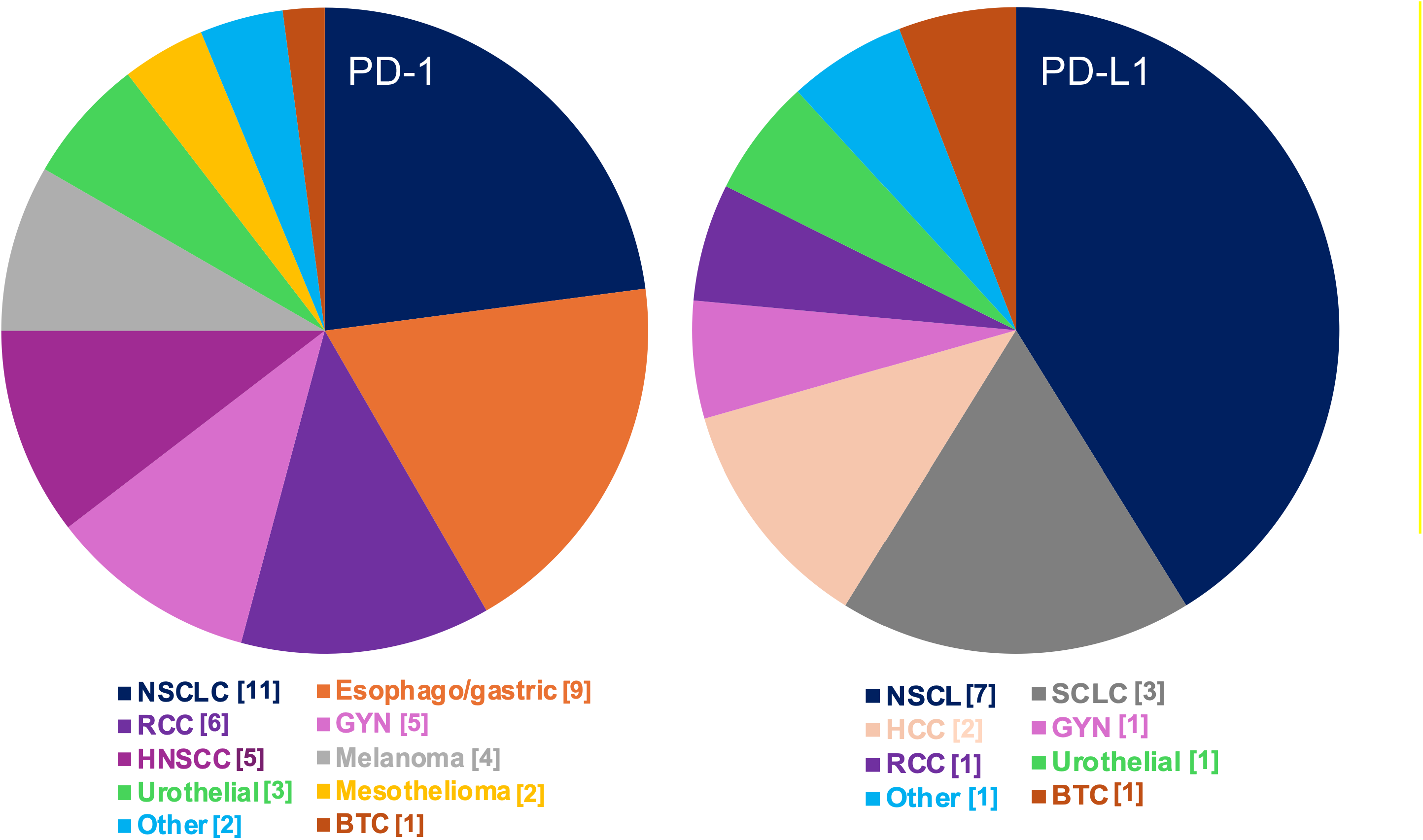
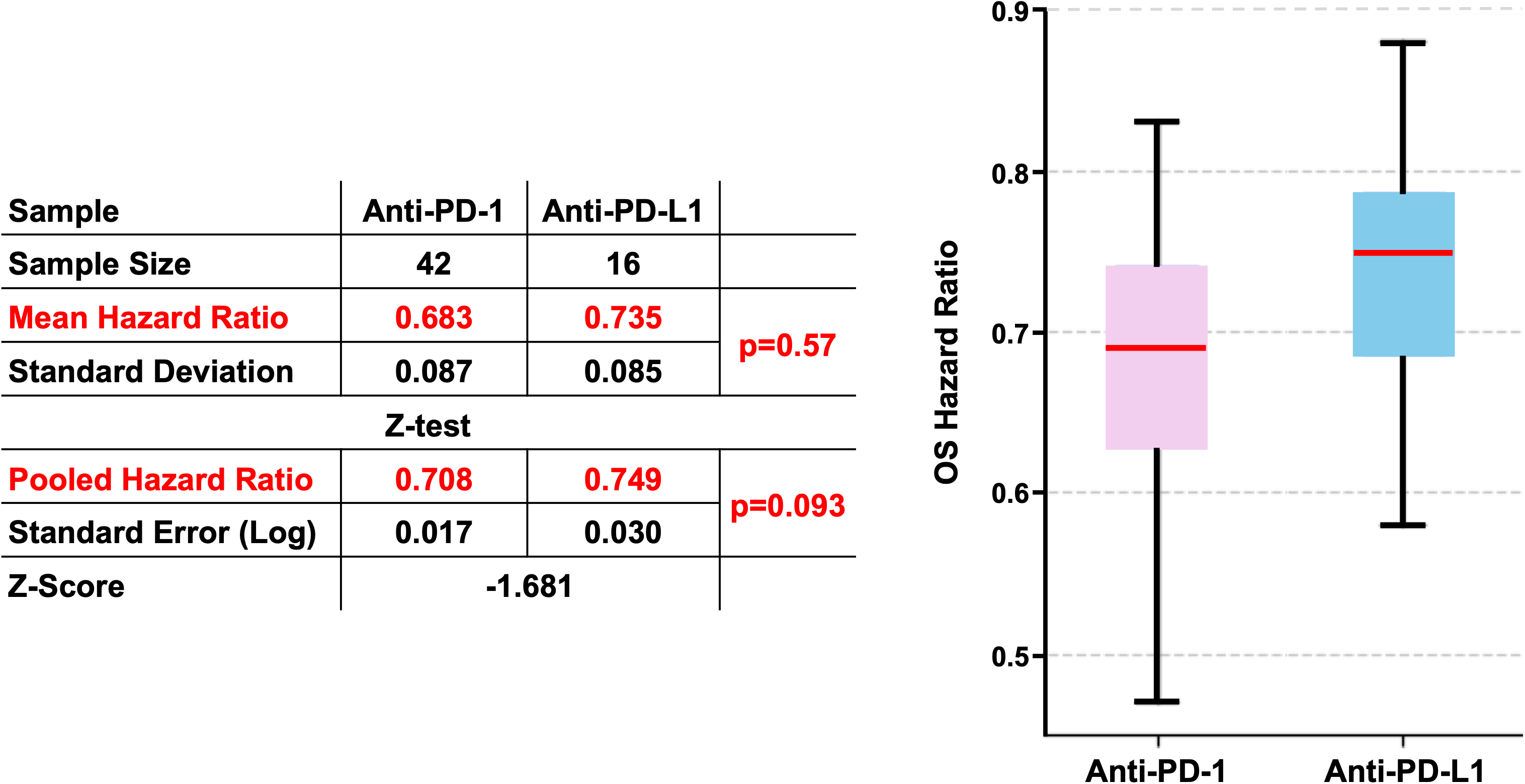
[A] Tumors treated with anti-PD-1 and anti PD-L1 emerge as similar. [**B**] Statistically the OS hazard ratios for anti-PD-1 and anti PD-L1 agents overlap.

## DISCUSSION

### Duration of approval process

While the era of immunotherapy approvals began with ipilimumab on March 25, 2011, it would be September 4, 2014, three-and-one-half years later, before the next regulatory approval for an immunotherapy was granted to pembrolizumab for unresectable/advanced melanoma. After that the approvals came rolling in. Our analysis begins with this pembrolizumab approval by the FDA and finds in the ensuing three to four years a more rapid pace of immunotherapy approvals with medians of 869 - 1144 days from time 0 defined as the study start date in Clintrials.gov to the approval letter, a pace that slowed and in the 2019 to 2024 period had approvals occurring a remarkably/unacceptably long median of 1338 – 2079 days. The rapid pace between 2014 and 2018, coincided with the exuberance over immunotherapy. Overall survival was an approval endpoint in only 13 (30%) of the 43 approvals between 2014 and 2018 with ORR an approval endpoint in 35 (81%) of the 43 approvals, with 26 (60%) of the 43 granted as accelerated approvals, including 18 based on single arm, non-randomized studies. Four of these approvals were later rescinded^8^ [Me. For the 99 indications approved staring in 2019, 45 (45%) and 60 (61%) were approved based on OS or an event-free endpoint and 60 (60%) of the 99 based on an ORR endpoint. Still 24/60 (40%) of those approved based on an ORR endpoint were accelerated approvals.

### Surrogate endpoints

Across all approvals, endpoints described as surrogates – including progression-, disease-, event- and relapse-free survivals (PFS, DFS, EFS, RFS, henceforth FS endpoints) as well as overall response rates and duration of responses (ORR, DOR) – were 74% and 74% of the primary or co-primary endpoints in clinical trials of immunotherapies and non-immunotherapies, respectively. These results coincide with previous studies reporting increasing use of surrogates such as PFS [Kemp and Prasad, 2017]. Advocates for the use of surrogate endpoints have argued they can often be measured sooner than OS, speed up readouts of efficacy and accelerate regulatory approvals. However, the deployment of a surrogate requires a rigorous approach to their assessment and should not exceed guidance given by regulatory bodies since they can also be potentially misleading^2, 9–14^. While indeed surrogate endpoints can often be measured sooner than OS, our data show comparable durations to approval for both immunotherapy and non-immunotherapy trials with an OS endpoint as those with FS endpoints; indeed, even comparable to those with an OOR/DOR endpoint although these latter endpoints had much smaller trial size. Median time to approvals of 1446 days for immunotherapy trials was slightly less than the median time of 1685 days for those that assessed non-immunotherapies, influenced by the faster approvals during the initial few years, but have since then grown closer and for non- immunotherapies have not changed meaningfully over more than a decade.

### Modest PFS and OS gains

Have we made progress compared to a similar analysis performed in 2014 of 71 FDA drug approvals when the median gains in PFS and OS were 2.5 and 2.1 months respectively?^15^ The data support a qualified yes as the answer, but the very modest gains underscore what every oncologist knows – cancer remains a formidable foe. Median PFS gains of 3.7 months, and median OS gains of 3.2 months for both immunotherapies and non-immunotherapies combined amount to an improvement of slightly more than one month. However, a direct comparison cannot be made. The current dataset shows a higher fraction of front-line therapies than the previous analysis, especially with immunotherapies where many moved rapidly to the front-line setting and studies with slight variation in design or with a similar design but examining a different checkpoint inhibitor emulated successful previous trials and contributed to the higher median values. Even more important, in the previous analysis we bemoaned the emergence of me-too therapies and over time this has blossomed as evidenced by the many anti-PD-1 and anti-PD-L1 agents and the plethora of therapies for tumors harboring EGFR and ALK alterations, amongst others. Trials with very similar therapies conducted to emulate a previously successful trial may have exaggerated the actual benefit.

A “check” on FDA approvals during this period is provided by the approvals of the EMA. In looking at the results **in Table 3** one can see the concerns of the EMA, that many would agree with. The 37 trials make it clear that the EMA feels less comfortable than the FDA in approving single arm trials or trials where ORR/DOR are the approval endpoints. Additionally, it has disagreed with the FDA in seven randomized trials that did not achieve statistically superior overall survivals with the experimental intervention.

**Table 3.** Therapies Approved by US Food and Drug Administration [USFDA] but not the European Medicines Agency [EMA].

| Drug | Disease | Study / Trial | Endpoint | Accelerated/<br>Full Approval | Annual<br>Incidence | Study<br>Size | Days to<br>Approval | ORR [CR]<br>[Duration of<br>benefit] | Approval<br>Date | NCT |
| --- | --- | --- | --- | --- | --- | --- | --- | --- | --- | --- |
|  |  |  |  |  |  |  |  | PFS HR<br>OS HR |  |  |
| Single Arm Trials 1<sup>st</sup> Line |  |  |  |  |  |  |  |  |  |  |
| Iobenguane <sup>131</sup>I | Pheochromocytoma and paraganglioma | IB12B | ORR | No/No | 50-800 | 68 | 3,313 | 22% [0%]<br>[53% 6 mos] | 07/30/18 | NCT00874614 |
| Pembrolizumab | Merkel carcinoma | KEYNOTE-017 | ORR/DOR | Yes/No | 2,000-3,000 | 50 | 1,485 | 56% [24%]<br>[98% 6 mos] | 12/19/18 | NCT02267603 |
| Pexidartinib | Tenosynovial giant cell tumor | ENLIVEN | ORR/DOR | No/No | 4,000 | 120 | 1,544 | 38% [15%]<br>[98% 6 mos] | 08/02/19 | NCT02371369 |
| Tazemetostat | Epithelioid Sarcoma | EZH-202 | ORR | Yes/No | 150 | 250 | 1,493 | 15% [1.6%]<br>[67% 6 mos] | 01/23/20 | NCT02601950 |
| Mitomycin gel | Low grade upper tract urothelial cancer | OLYMPUS | ORR/DOR | No/No | 6,000-8,000 | 71 | 1,107 | 58% [58%]<br>[27% 6 mos] | 04/15/20 | NCT02793128 |
| Sirolimus protein-bound particles inject suspension | PEComa | AMPECT | ORR/DOR | No/No | <350 | 31 | 2,244 | 39% [6%]<br>[67% 12 mos] | 11/22/21 | NCT02494570 |
| Single Arm Trials 2<sup>nd</sup> Line |  |  |  |  |  |  |  |  |  |  |
| Copanlisib | Follicular Lymphoma | CHRONOS-1 | ORR/DOR | Yes/WD<br>[WD globally] | 15,000 | 104 | 1,865 | 58.7% [14%]<br>[12.2 mos] | 09/14/17 | NCT01660451 |
| Pembrolizumab | PMLBCL | KEYNOTE 170 | ORR/DOR | Yes/No | 140 | 53 | 924 | 45% [11%]<br>[NA] | 06/13/18 | NCT02576990 |
| Pembrolizumab | SCLC | KEYNOTE-158G<br>KEYNOTE-028C | ORR/DOR | Yes/WD | 30,000 | 83 | 1,278/1,933 | 19% [2%]<br>[63% 12 mos] | 06/17/19 | NCT02628067<br>NCT02054806 |
| Pomalidomide | Kaposi Sarcoma HIV Pos<br>[Restricted/Compassionate] | 12-C-0047 | ORR/DOR | Yes/No | 2,500 | 28 | 3,047 | 67% [NA]<br>[12.5 mos] | 05/14/20 | NCT01495598 |
|  | Kaposi Sarcoma HIV Neg<br>[Restricted/Compassionate] |  |  |  |  |  |  | 80% [NA]<br>[10.5 mos] |  |  |
| Rucaparib + GnRH analog or prior bilateral orchiectomy | mCRPC + BRCA alteration | TRITON2 | ORR/DOR | Yes/No | 12,000-35,000 | 115 | 1,185 | 44%<br>[56% 6 mos] | 05/15/20 | NCT02952534 |
| Tazemetostat | Follicular Lymphoma + EZH2 mutation | E7438-G000-101 | ORR/DOR | Yes/WD<br>[WD globally] | 3,000-4,500 | 42 | 2,562 | 69% [12%]<br>[10.9 mos] | 06/18/20 | NCT01897571 |
| Selinexor | DLBCL | SADAL | ORR/DOR | Yes/No | 25,000-32,000 | 134 | 2,090 | 29% [13%]<br>[38% 6 mos] | 06/22/20 | NCT02227251 |
| Belantamab mafodotin-blmf | Multiple Myeloma | DREAMM-2 | ORR/DOR | Yes/WD | 36,000 | 221 | 779 | 31% [NA]<br>[73% 6 mos] | 08/05/20 | NCT03525678 |
| Pralsetinib | RET-mut MTC prior RX | ARROW | ORR/DOR | Yes/WD | 820-1,230 | 55 | 1,355 | 60% [NA]<br>[79% 6 mos] | 12/01/20 | NCT03037385 |
|  | RET-mut MTC no prior Rx |  |  |  |  | 29 |  | 66% [NA]<br>[84% 6 mos] |  |  |
|  | RET-fusion+ thyroid cancer |  |  |  |  | 9 |  | 89% [NA] |  |  |
|  |  |  |  |  |  |  |  | [100% 6 mos] |  |  |
| Umbralisib | Marginal Zone Lymphoma | UTX-TGR-205 | ORR/DOR | Yes/WD<br>[WD globally] | 8,000 | 69 | 1,717 | 49% [16%]<br>[NE] | 02/05/21 | NCT02793583 |
|  | Follicular Lymphoma |  |  |  | 15,000 | 117 |  | 43% [3%]<br>[11.1 mos] |  |  |
| Sacituzumab govitecan | Locally advanced or metastatic urothelial cancer | TROPHY-U-01 | ORR/DOR | Yes/WD | 12,000 | 112 | 1,247 | 27.7% [5.4%]<br>[7.2 mos] | 04/13/21 | NCT03547973 |
| Infigratinib | LA / metastatic CCA + FGFR fusion/rearrangement | CBGJ398X2204 | ORR/DOR | Yes/WD | 2,000-3,000 | 108 | 2,501 | 23% [1%]<br>[5 mos] | 05/28/21 | NCT02150967 |
| Mobocertinib | NSCLC / EGFR Exon 20 insertions | Study 101 | ORR/DOR | Yes/WD<br>[WD globally] | 20,000 | 114 | 1,917 | 28% [NA]<br>[17.5 mos] | 09/15/21 | NCT02716116 |
| Melphalan for Injection/Hepatic Delivery | Uveal melanoma | FOCUS | ORR/DOR | No/No | 1,700-2,500 | 91 | 2,751 | 36.3%<br>[14 mos] | 08/14/23 | NCT02678572 |
| Lifileucel | Melanoma | C-144-01 | ORR/DOR | Yes/No | 100,000 | 73 | 3,090 | 31.5% [NA]<br>{NE} | 02/16/24 | NCT02360579 |
| Tovorafenib | Low grade glioma / BRAF fusion / rearrangement or BRAF V600Emut | FIREFLY-1 | ORR/DOR | Yes/No | 3,400 | 76 | 1,097 | 51% [NA]<br>[13.8 mos] | 04/23/24 | NCT04775485 |
| Lisocabtagene maraleucel | Mantle cell lymphoma | TRANSCEND-MCL | ORR/DOR | No/No | 4,000 | 68 | 1,401 | 85.3% [67.6%]<br>[13.3 mos] | 05/30/24 | NCT02631044 |
| Afamitresgene autoleucel<sup>A</sup> | Synovial Sarcoma | SPEARHEAD-1 | ORR/DOR | Yes/No | 800-1000 | 52 | 1,816 | 43.2% [NA]<br>[6 mos] | 08/02/24 | NCT04044768 |
| Zenocutuzumab-zbco | NRG1 fusion NSCLC | eNRGy | ORR/DOR | Yes/No | <1,000<br>330 | 64 | 3,625 | 33% [NA]<br>[7.4 mos] | 12/04/24 | NCT02912949 |
|  | NRG1 fusion pancreatic cancer |  |  |  |  |  |  | 40% [NA]<br>[3.7-16.6 mos] |  |  |
| Cosibelimab-ipdl | Metastatic cutaneous squamous cell carcinoma | CK-301-101 | ORR/DOR | No/No | 30,000 | 78<br>31 | 2641 | 47% [NA]<br>{NR} | 12/13/24 | NCT03212404 |
|  | Locally advanced cutaneous squamous cell carcinoma |  |  |  |  |  |  | 48% [NA]<br>[17.7 mos] |  |  |
| Randomized Trials 1<sup>st</sup> Line |  |  |  |  |  |  |  |  |  |  |
| Necitumumab + gemcitabine/cisplatin | Squamous NSCLC | SQUIRE | PFS/OS | No/No | 66,000 | 1093 | 2,147 | 0.85 [0.74, 0.98]<br>0.84 [0.74, 0.96]<br>[XO Not Allowed] | 11/24/15 | NCT00981058 |
| Cabozantinib | Renal cell carcinoma | CABOSUN | PFS | No/No | 18,000-24,000 | 157 | 1595 | 0.48; [0.31, 0.74]<br>**0.80 [0.50, 1.26]**<br>[XO Not Allowed] | 12/19/17 | NCT01835158 |
| Atezolizumab + cobimetinib/vemurafenib | BRAF V600mut unresectable or metastatic melanoma | IMspire150 | PFS | No/No | 40,000-60,000 | 514 | 1,294 | 0.78 [0.63, 0.97]<br>**0.84 [0.67, 1.05]**<br>[XO Not Allowed; but post protocol Rx] | 07/30/20 | NCT02908672 |
| Dabrafenib + Trametinib | BRAF V600E mut low-grade glioma | CDRB436G2201 | ORR/PFS | No/No | 28-70 | 110 | 1,904 | 46.6% vs. 10.8%<br>0.31 [0.17, 0.55]<br>NE | 03/16/23 | NCT02684058 |
| Acalabrutinib + bendamustine/rituximab | Mantle Cell Lymphoma | ECHO | PFS | No/No | 4,000 | 598 | 2,843 | 0.73 [0.57, 0.94]<br>**0.86 [0.65, 1.13]**<br>[XO Allowed. | 01/16/25 | NCT02972840 |
|  |  |  |  |  |  |  |  | 20 + 5 = 25%] |  |  |
| **Randomized Trials 2<sup>nd</sup> Line** |  |  |  |  |  |  |  |  |  |  |
| **Cabozantinib** | Advanced renal cell carcinoma | METEOR | PFS/OS | **No/No** | 18,000-24,000 | 375 [658] | 1059 | 0.58 [0.45, 0.74]<br>0.66 [0.53, 0.83] | 04/25/16 | NCT01865747 |
| **Margetuximab-cmkb** | HER2+ Breast Cancer | SOPHIA | PFS/OS | **No/No** | 63,000-79,000 | 536 | 1,941 | 0.76 [0.59, 0.98]<br>**0.95 [0.77, 1.17]**<br>[XO Not Allowed] | 12/16/20 | NCT02492711 |
| **Inavolisib + palbociclib/fulvestrant** | PIK3CA-mut breast Cancer | INAVO120 | ORR/PFS/OS | **No/No** | 110,000-126,000 | 325 | 1,716 | 0.43 [0.32, 0.59]<br>0.67 [0.48, 0.94]<br>[XO Not Allowed.<br>8.5-10% other PI3Ki] | 10/10/24 | NCT04191499 |
| **Ensartinib** | ALK+ NSCLC | eXALT3 | PFS/OS | **No/No** | 5,800-10,000 | 290 | 3,122 | 0.56 [0.40, 0.79]<br>**0.88 [0.63, 1.23]**<br>[XO Not Allowed] | 12/18/24 | NCT02767804 |
| **Sotorasib + panitumumab** | Colorectal cancer / KRAS G12C mutation | CodeBreak 300 | PFS/[OS] | **No/No** | 5,500 | 160 | 1,003 | 0.48 [0.30, 0.78]<br>**0.70 [0.41, 1.18]**<br>[XO Not Allowed] | 01/16/25 | NCT05198934 |
| **Datopotamab deruxtecandlnk** | HR+/Her-2- Breast Cancer | TROPION-Breast01 | PFS/OS/<br>ORR | **No/No** | 220,000-237,000 | 732 | 1,187 | 0.63 [0.52, 0.76]<br>**1.01 [0.83, 1.22]**<br>[XO Not Allowed] | 01/17/25 | NCT05104866 |

Accelerated / Full, accelerated approval / full approval;
Abbreviations: ALK, anaplastic lymphoma kinase; AML, acute myelocytic leukemia; BRAF, proto-oncogene BRAF; CCA, cholangiocarcinoma; DOR, duration of response; FL, follicular lymphoma; Her2, human epidermal growth factor receptor-2; HR, hormone receptor; HRD, homologous recombination deficiency; IDH1, isocitrate dehydrogenase 1; KRAS, Kirsten rat sarcoma virus oncogene homolog; LACCA, locally advanced cholangiocarcinoma; MCL, mantle cell lymphoma; MCCA, metastatic cholangiocarcinoma; MZL, marginal zone lymphoma; MTC, medullary thyroid cancer; N, no; NSCLC, Non-small cell lung cancer; ORR, overall response rate; OS, overall survival; PEComa, perivascular epithelioid cell tumor; PFS, progression-free survival; PIK3CA, phosphatidylinositol-4,5-bisphosphate 3-kinase catalytic subunit alpha; PMLBCL, primary mediastinal large b-cell lymphoma; PHEO/PPGL, pheochromocytoma and paraganglioma; RCC, renal cell carcinoma; rrMDS, relapsed or refractory myelodysplastic syndromes; STS, soft tissue sarcoma; **WD, withdrawn**; XO, crossover; Y, yes.
^Tumors HLA-A\*02:01P, -A\*02:02P, -A\*02:03P, or -A\*02:06P positive and whose tumor expresses the MAGE-A4 antigen

### Translation of benefit from FS to OS

Beyond the concerns of the EMA, our analysis comparing immunotherapy and non-immunotherapy data yielded interesting observations. One that stands out emerged from looking at the HRs of the FS and OS analyses that revealed that the FS endpoint statistical advantage for the non-immunotherapy trials disappeared in the OS analysis. This marked decay in translation of the FS benefit to OS with non-immunotherapies [61%] can be seen as concerning but may be in part explained by toxic therapies and informative censoring with a median close to 30% of the data censored in both arms [not shown]. This decay contrasted with our observation in immunotherapy trials, in which the FS benefit *did* translate to OS [99%]. Note that this does not imply OS is a better endpoint for immunotherapies, nor that with time the benefit improves. It simply indicates the FS benefit was largely translated to the OS endpoint.

Despite the exuberance that the coming of immunotherapies generated, and the paradigm-shift that they created for oncology, in the end their performance has been challenged by the non-immunotherapy offerings, primarily targeted therapies, with the evolution of better agents for older targets – EGFR and ALK as examples – as well as the brisk development of diverse options for new targets (MET, RET, NTRK, CDK4/6 and PSMA as examples), and the development of novel approaches for these targets in the antibody drug conjugates. The gains not in surrogates but in overall survival with several targeted therapies were enormously gratifying and gives one hope that as other targets are found, tweaking of the first generation can lead to very effective agents.

### Endpoints for drug approval

What does the analysis tell us should be done? We would argue, as have many others, that our analysis highlights the need for a major overhaul of the drug approval process^16–20^. While regulatory agencies acknowledge the need to modernize the approval process with tissue agnostic approvals as one example^21,22^, in fact the major metrics for approvals have not changed in a meaningful way in over four decades, although the frequency of their use has varied^23–28^. Response rate, OS and FS endpoint surrogates such as progression/disease/relapse/event-free survival have been the mainstay of drug approvals, with the latter arriving in the mid 1980’s and widely accepted a few years later^26,29,30^. We believe nearly everyone would agree a median time *for pivotal trial conduct* of more than five years [1708 – 2357 days] cannot continue as the acceptable status quo for trials that score regulatory approvals. Especially given the years of research and early phase clinical trials that precede the launch of the registration trial. While a duration of more than five years might be seen as acceptable were the primary endpoint OS requiring time to mature, this has occurred even as “surrogate endpoints” surged with the data pointing to comparable times to approvals across endpoints [**Figure 3**]. We recognize decisions as to which endpoint is pursued may be influenced by the disease and its “natural history – e.g., choosing an “FS” endpoint for a disease such as a neuroendocrine cancer and OS for a more rapidly lethal disease such as pancreatic cancer. However, the unacceptably long duration to approvals is in part being driven by the focus on statistical significance for often-marginal results, often requiring trials with large number of enrollees – use of the CDK4 inhibitors abemaciclib and ribociclib as adjuvant therapies, as examples^31^.

### Too many trials

Additionally, the focus on statistical significance highlights an important point that many find uncomfortable: acknowledging that drug trials and drug approvals today are driven not by the need for novel cancer therapeutics but rather by financial/economic factors, specifically pharmaceutical company revenue. This unfortunately is an inescapable truth. While as a society we derive benefits from this economic endeavor, it is far from the optimal path to drug development with the immunotherapy era a glaring example. As evidence consider the 22 immunotherapy trials in non-small cell lung cancer that enrolled 13,325 patients over more than a decade and served as registration trials. Together they provide sound evidence that indeed checkpoint inhibitors are effective in NSCLC, across the spectrum of the disease. The redundancy of the trials, as well as their often-minimal differences were not accidental but rather carefully planned strategies by pharmaceutical companies, excellent examples of risk-averse enterprises seeking a niche they could exploit for revenue. The HRs of the trials are remarkably similar, because the drugs are remarkably similar and the disease the same. We can argue without doubt that we did not need 22 trials and 13,325 patients to establish the efficacy of checkpoint inhibitors in NSCLC. Its efficacy was never in doubt after the first several hundred patients had been treated. A fraction of 13,325 would have reached the same conclusion but instead, we sit now on a mountain of replicative data, minimally tweaked from trial to trial and unfortunately lack critical information that has not been pursued – the optimal dose, the optimal interval of treatment and the optimal number of cycles needed. Very compelling or limited data exists that much lower doses can be effective^32–39^, that the interval between administrations can be longer^40,41^ and that many fewer cycles of therapy are needed^42,43^. As regards comparability of the drugs we would note here that a crude analysis of the data in **Table 2** that many could argue is flawed but that simply compares data from trials that enrolled very similar patients finds the anti-PD-L1 antibodies with a pooled HR for OS of 0.75 [0.71, 0.79] and that of the anti-PD-1 agents with a pooled HR for OS of 0.71 [0.69, 0.73], statistically similar. A sensitivity analysis confirmed the results are highly stable, and the conclusion of similarity is robust. Whether we looked at the raw averages, the weighted Z-test, or a sensitivity analysis excluding outliers, the statistical conclusion remained the same, the groups are similar.

### Excess patients in control arms

We would argue novel drug development strategies should aim to markedly reduce trial size, which would in turn reduce trial duration and get truly effective therapies to patients sooner. Large trial size inevitably means the same for the control arm. In NSCLC, novel strategies to assess efficacy and analyses validating the use of “external or synthetic controls” could have kept many of the 5,177 patients who enrolled on the reference arm of the aforementioned 22 trials from receiving the inferior regimen^44^. Advocacy for eliminating or markedly reducing the control arm once it is shown to emulate existing “external or synthetic controls” has many advocates – with small cell lung cancer, pancreatic cancer, diffuse large B-cell lymphoma and now NSCLC as prime examples of tumors where countless trials using the same control arm were conducted over many years^45–51^.

### Accelerated approval as a solution

Note here that the answer does not lie in increasing the number of accelerated approvals. Despite study sizes of 110 for accelerated approvals, less than one half of the 489-patient size of “regular approvals”, accelerated approvals were not accomplished in less time. Such approvals were nearly always applied to less common cancers and granted after a median of 4.14 years, unfortunately like the 4.45 years for full approvals. We previously stated that smaller studies should take less time; perhaps the emphasis on less common cancers undermined that promise. Efforts should also be made to give accelerated approval to novel therapies, not me-too options^15^, with meaningful magnitudes of benefit. Additional suggestions include agreeing on more stringent definitions of what constitutes an appropriate “surrogate” with endorsement only of “surrogates” that reproducibly lead to overall survival benefits^2,12,52–54^.

## Conclusion

While encouraging in many ways, the results highlight the remaining challenges. Although curative therapies are what the public often thinks we are pursuing and those who enroll on clinical trials are often expecting, in fact neither the trajectory of the data nor the design of clinical trials supports such a view. Prolongation of life is the more realistic goal of drug development and clinical trials, and the metric endorsed by regulatory agencies that vet these efforts.

However, the future looks promising, with an increasing number of indications achieving meaningful prolongations of life, allowing us to tell an increasing number of patients that while it is still difficult to achieve cures, meaningful prolongation of their lives may be possible, and this may mean the next therapy they receive could be a very novel one that has yet to be developed.

## Data Availability

All data produced in the present study are available upon reasonable request to the authors.

https://www.fda.gov/drugs/resources-information-approved-drugs/oncology-cancerhematologic-malignancies-approval-notifications

## REFERENCES

1. Phan GQ, Yang JC, Sherry RM, Hwu P, Topalian SL, Schwartzentruber DJ, et al. Cancer regression and autoimmunity induced by cytotoxic T lymphocyte-associated antigen 4 blockade in patients with metastatic melanoma. Proc Natl Acad Sci U S A. 2003; 100:8372–7. doi: 10.1073/pnas.1533209100. PMID: 12826605.

2. Kemp R, Prasad V. Surrogate endpoints in oncology: when are they acceptable for regulatory and clinical decisions, and are they currently overused? BMC Med. 2017; 15:134. doi: 10.1186/s12916-017-0902-9. PMID: 28728605.

3. Gyawali B, Rome BN, Kesselheim AS. Regulatory and clinical consequences of negative confirmatory trials of accelerated approval cancer drugs: retrospective observational study. BMJ. 2021; 374:n1959. doi: 10.1136/bmj.n1959. PMID: 34497044

4. Gyawali B, Hey SP, Kesselheim AS. Assessment of the Clinical Benefit of Cancer Drugs Receiving Accelerated Approval. JAMA Intern Med. 2019; 179:906–913. doi: 10.1001/jamainternmed.2019.0462. PMID: 31135808.

5. Liu ITT, Kesselheim AS, Cliff ERS. Lessons from withdrawn accelerated approvals in oncology. JAMA. 2024; 331:1471–1479. doi:10.1001/jama.2024.2396. PMID: 38583175.

6. Project Confirm resources. https://www.fda.gov/about-fda/oncology-center-excellence/project-confirm.

7. Accelerated Approval Program resources. https://www.fda.gov/drugs/nda-and-bla-approvals/accelerated-approval-program

8. Mellgard GS, Fojo T, Bates SE. Lessons from withdrawn accelerated approvals in oncology. Nat Cancer. 2024; 5:211–215. doi: 10.1038/s43018-023-00696-8. PMID: 38291305.

9. 9. Surrogate Endpoints resource. https://www.cancer.gov/publications/dictionaries/cancer-terms/def/surrogate-endpoint

10. Ciani O, Buyse M, Drummond M, Rasi G, Saad ED, Taylor RS. Time to Review the Role of Surrogate End Points in Health Policy: State of the Art and the Way Forward. Value Health. 2017; 20:487–495. doi: 10.1016/j.jval.2016.10.011. PMID: 28292495.

11. Belin L, Tan A, De Rycke, Y. Dechartres A. Progression-free survival as a surrogate for overall survival in oncology trials: a methodological systematic review. Br. J. Cancer. 2020. doi: 10/1038/s41416-202-0805-y. PMID: 32214230.

12. Mittal A, Kim MS, Dunn S, Wright K, Gyawali B. Frequently asked questions on surrogate endpoints in oncology-opportunities, pitfalls, and the way forward. EClinicalMedicine. 2024; 76:102824. doi: 10.1016/j.eclinm.2024.102824. PMID: 39764569.

13. Jeng LJB, Siegel J. Surrogate Endpoints in Regulatory Decision-Making Clin Transl Sci. 2025; 18:e70445. doi: 10.1111/cts.70445. PMID: 41414877.

14. Chen EY, Joshi SK, Tran A, Prasad V. Estimation of Study Time Reduction Using Surrogate End Points Rather Than Overall Survival in Oncology clinical Trials. JAMA Intern Med. 2019a; 179:642–647. doi: 10.1001/jamainternmed.2018.8351. PMID: 30933235.

15. Fojo T, Mailankody S, Lo A. Unintended consequences of expensive cancer therapeutics—the pursuit of marginal indications and a me-too mentality that stifles innovation and creativity: the John Conley Lecture. JAMA Otolaryngol Head Neck Surg. 2014; 140:1225–1236. doi: 10.1001/jamaoto.2014.1570. PMID: 25068501.

16. Skydel JJ, Luxkaranayagam AT, Dhruva SS, Ross JS, Wallach JD. Analysis of Postapproval Clinical Trials of Therapeutics Approved by the US Food and Drug Administration Without Clinical Postmarketing Requirements or Commitments. JAMA Netw Open. 2019; 2:e193410. doi: 10.1001/jamanetworkopen.2019.3410. PMID: 31074812.

17. Zhang AD, Puthumana J, Downing NS, Shah ND, Krumholz HM, Ross JS. Assessment of Clinical Trials Supporting US Food and Drug Administration Approval of Novel Therapeutic Agents, 1995-2017. JAMA Netw Open. 2020; 3:e203284. doi: 10.1001/jamanetworkopen.2020.3284. PMID: 32315070.

18. Mitra-Majumdar M, Gunter SJ, Kesselheim AS, Brown BL, Joyce KW, Ross M, Pham C, Avorn J, Darrow JJ. Analysis of Supportive Evidence for US Food and Drug Administration Approvals of Novel Drugs in 2020. JAMA Netw Open. 2022; 5:e2212454. doi: 10.1001/jamanetworkopen.2022.12454. PMID: 35579897.

19. Dhruva SS, Darrow JJ, Kesselheim AS, Redberg RF. Strategies to Manage Drugs and Devices Approved Based on Limited Evidence: Results of a Modified Delphi Panel. Clin Pharmacol Ther. 2022; 111:1307–1314. doi: 10.1002/cpt.2583. PMID: 35292958.

20. Koole SN, Huisman AH, Timmers L, Westgeest HM, van Breugel E, Sonke GS, et al. Lessons learned from postmarketing withdrawals of expedited approvals for oncology drug indications. Lancet Oncol. 2024; 25:e126–e135. doi: 10.1016/S1470-2045(23)00592-2. PMID: 38423058.

21. Thein KZ, Myat YM, Park BS, Panigrahi K, Kummar S. Target-Driven Tissue-Agnostic Drug Approvals-A New Path of Drug Development. Cancers (Basel). 2024; 16:2529. doi: 10.3390/cancers16142529. PMID: 39061168.

22. Sledge GW Jr, Yoshino T, Xiu J, Helmstetter A, Ribeiro JR, Klimov S, et al. Real-world evidence provides clinical insights into tissue-agnostic therapeutic approvals Nat Commun. 2025; 16:2646. doi: 10.1038/s41467-025-57941-0. PMID: 40102447.

23. Johnson JR, Williams G, Pazdur R.J. End points and United States Food and Drug Administration approval of oncology drugs. Clin Oncol. 2003; 21:1404–11. doi: 10.1200/JCO.2003.08.072. PMID: 12663734.

24. Wilson MK, Karakasis K, Oza AM. Outcomes and endpoints in trials of cancer treatment: the past, present, and future. Lancet Oncol. 2015;16:e32–42. doi: 10.1016/S1470-2045(14)70375-4. PMID: 25638553.

25. Chen EY, Raghunathan V, Prasad V. An Overview of Cancer Drugs Approved by the US Food and Drug Administration Based on the Surrogate End Point of Response Rate. JAMA Intern Med. 2019b; 179:915–921. doi: 10.1001/jamainternmed.2019.0583. PMID: 31135822.

26. Chitkara A, Manoj P Rai, Thawani R, Chen EY. Recent analysis of frequency of surrogate endpoints used in oncology clinical trials 2006-2022. J Clin Oncol 41, 2023 (suppl 16; abstr e13658).

27. Sharma R, Gulati A, Chopra K. Accelerated approvals in oncology: Trial design strategies and insights driving successful regulatory outcomes across three decades. Int J Cancer. 2026; 158:2853–2865. doi: 10.1002/ijc.70317. PMID: 41460153.

28. Shahnam A, DiConza K, Jasnani S, Desir-Camille R, Rosenbaum E, Babatunde O, et al. Objective response rate and progression-free survival as surrogates for overall survival treatment effect: A meta-analysis across diverse tumour groups and contemporary therapies. Cancer Med. 2026; 15:e72066. doi: 10.1002/cam4.72066. PMID: 42324568.

29. Raphael MJ, Robinson A, Booth CM, O’Donnell J, Palmer M, Eisenhauer E, et al. The Value of Progression-Free Survival as a Treatment End Point Among Patients with Advanced Cancer. A Systematic Review and Qualitative Assessment of the Literature. JAMA Oncol. 2019; 5:1779–1789. doi: 10.1001/jamaoncol.2019.3338. PMID: 31556921.

30. Sherry AD, Miller AM, Parlapalli JP, Kupferman GS, Beck EJ, McDonald J, et al. Overall Survival and Quality-of-Life Superiority in Modern Phase 3 Oncology Trials. A Meta-Epidemiological Analysis. JAMA Oncol. 2025; 11:718–724. doi: 10.1001/jamaoncol.2025.1002. PMID: 40451185.

31. Tannock IF, Khan QJ, Fojo T. Why We Do Not Recommend That Women With Breast Cancer Receive Adjuvant Treatment with a CDK4/6 Inhibitor. J Clin Oncol. 2025; 43:2456–2460. doi: 10.1200/JCO-24-02683. PMID: 40228181.

32. Chatterjee M, Turner DC, Felip E, Lena H, Cappuzzo F, Horn L, et al. Systematic evaluation of pembrolizumab dosing in patients with advanced non-small-cell lung cancer. Ann Oncol. 2016; 27:1291–8. doi: 10.1093/annonc/mdw174. PMID: 27117531.

33. Low JL, Huang Y, Sooi K, Ang Y, Chan ZY, Spencer K, et al. Low dose pembrolizumab in the treatment of advanced non-small cell lung cancer. Int J Cancer. 2021; 149:169–176. doi: 10.1002/ijc.33534. PMID: 33634869.

34. Brahmer JR, Drake CG, Wollner I, Powderly JD, Picus J, Sharfman WH, et al. Phase I Study of Single-Agent Anti-Programmed Death-1 (MDX-1106) in Refractory Solid Tumors: Safety, Clinical Activity, Pharmacodynamics, and Immunologic Correlates. J Clin Oncol. 2023; 41:715–723. doi: 10.1200/JCO.22.02270. PMID: 36706735.

35. Qian Y, Yuan Wu Y, Qin J, Jin X. Efficacy and safety of low-dose versus standard-dose immune checkpoint inhibitor as monotherapy in patients with solid tumors: A meta-analysis and systematic review.2024. J Clin Oncol 42(23_suppl):188–188. DOI 10.12000/JCO.2024.42.23 suppl.23, abstract 188.

36. Remon J, Bortolot M, Bironzo P, Cortiula F, Menis J, Brandao M, et al. De-Escalation Strategies with Immune Checkpoint Blockers in Non-Small Cell Lung Cancer: Do We Already Have Enough Evidence? J Clin Oncol. 2025; 43:1148–1156. doi: 10.1200/JCO-24-02347. PMID: 39836933.

37. Sun J, Hao J, Li X, Xu L, Han T, Shi S, et al. A real-world study on the efficacy and safety of low-dose PD-1 monoclonal antibody alone or in combination as the first-line treatment for advanced non-small cell lung cancer. J Immunother Cancer. 2025; 13:e011622. doi: 10.1136/jitc-2025-011622. PMID: 40550562.

38. Jiménez-Labaig P, Mohamed F, Tan NJI, Sanna I, El Bairi K, Khan SZ, et al. Expanding access to cancer immunotherapy: A systematic review of low-dose PD-(L)1 inhibitor strategies. Eur J Cancer. 2025; 225:115564. doi: 10.1016/j.ejca.2025.115564. PMID: 40513286.

39. Noronha V, Patil V, Menon N, Shah M, Ostwal V, Ramaswamy A, et al. Efficacy and Safety of Ultra-Low-Dose Immunotherapy in Relapsed Refractory Solid Tumors: Phase III Superiority Randomized Trial (DELII). J Clin Oncol. 2026; 44:1083–1097. doi: 10.1200/JCO-25-01546. PMID: 41604598.

40. Hijmering-Kappelle LBM, Hiltermann TJN, Bensch F. Safety and Efficacy of Extended Interval Dosing for Immune Checkpoint Inhibitors in Non–Small Cell Lung Cancer During the COVID-19 Pandemic. Clin Lung Cancer. 2021. doi: 10.1016/j.cllc.2021.12.005. PMID: 35034861.

41. Cantini L, Paoloni F, Pecci F, Spagnolo F, Genova C, Tanda ET, et al. Safety of extended interval dosing immune checkpoint inhibitors: a multicenter cohort study. J Natl Cancer Inst. 2023; 115:796–804. doi: 10.1093/jnci/djad061. PMID: 37042716.

42. Chen Z, Li MSC, Lam WKJ. Optimal duration of adjuvant immunotherapy after neoadjuvant chemoimmunotherapy in resectable non-small cell lung cancer: a systematic review and meta-analysis. J Thorac Dis. 2026; 18:116. doi: 10.21037/jtd-2025-1389. PMID: 41816378.

43. Bazarbachi AH, Magrini N, Aziz Z, Fojo T. Evidence for a reduction in number of cycles of immune checkpoint inhibitors. Lancet Oncol. 2025; 26:9–11. doi: 10.1016/S1470-2045(24)00594-1. PMID: 39756447.

44. Mishra-Kalyani PS, Amiri Kordestani L, Rivera DR, Singh H, Ibrahim A, DeClaro RA, et al. External control arms in oncology: current use and future directions. Ann Oncol. 2022; 33:376–383. doi: 10.1016/j.annonc.2021.12.015. PMID: 35026413.

45. Yeh C, Bates SE. Two decades of research toward the treatment of locally advanced and metastatic pancreatic cancer: Remarkable effort and limited gain. Semin Oncol. 2021; 48:34–46. doi: 10.1053/j.seminoncol.2021.01.001. PMID: 33712267.

46. Azizi Z, Zheng C, Mosquera L, Pilote L, El Emam K; GOING-FWD Collaborators. Can synthetic data be a proxy for real clinical trial data? a validation study. BMJ Open. 2021; 11:e043497. doi:10.1136/bmjopen-2020-043497. PMID: 33863713.

47. Popat S, Liu SV, Scheuer N, Hsu GG, Lockhart A, Ramagopalan SV, et al. Addressing challenges with real-world synthetic control arms to demonstrate the comparative effectiveness of Pralsetinib in non-small cell lung cancer. Nat Commun. 2022; 13:3500. doi:10.1038/s41467-022-30908-1. PMID: 35715405.

48. Banerjee R, Midha S, Kelkar AH, Goodman A, Prasad V, Mohyuddin GR. Synthetic control arms in studies of multiple myeloma and diffuse large B-cell lymphoma. Br J Haematol. 2022; 196:1274–1277. doi:10.1111/bjh.17945. PMID: 34750805.

49. Piron B, Rodrigo L, Tessoulin B, Gastinne T, Dubruille V, Letailleur V, et al. Synthetic control arm from mixed clinical trials and real-world data from the LYSA group for untreated diffuse large B-cell lymphoma patients aged over 80 years: a *bona fide s*trategy for innovative clinical trials. Clin Lymphoma Myeloma Leuk. 2026; 26:115–122.e6. doi: 10.1016/j.clml.2025.10.024. PMID: 41274822.

50. Zhou M, Fojo AT, Schwartz LH, Bates SE, Blagoev KB. Cancer interception during treatment: using growth kinetics to create a continuous variable for assessing disease response. Oncologist. 2025; 30:oyaf353. doi: 10.1093/oncolo/oyaf353. PMID: 41124020.

51. Malinou JN, Fan J, Cheng J, Gong Y, Shen YL, Larkins E. An FDA analysis of the association of tumor growth rate, overall survival, and progression-free survival in patients with metastatic non-small cell lung cancer Oncologist. 2026; 31:oyag009. doi: 10.1093/oncolo/oyag009. PMID: 41546388.

52. Begg CB. Justifying the Choice of Endpoints for Clinical Trials. J Natl Cancer Inst 2013; 105: 1594–1595. doi: 10.1093/jnci/djt289. PMID: 24108810.

53. Prasad V, Kim C, Burotto M and Vandross A. The Strength of Association Between Surrogate End Points and Survival in Oncology. A Systematic Review of Trial-Level Meta-analyses. JAMA Intern Med. 2015; 175:1389–1398. doi:10.1001/jamainternmed.2015.2829. PMID: 26098871.

54. Clinical Trial Considerations to Support Accelerated Approval of Oncology Therapeutics. March 2023. https://www.fda.gov/regulatory-information/search-fda-guidance-documents/clinical-trial-considerations-support-accelerated-approval-oncology-therapeutics.

